# Real-world epilepsy monitoring with ultra long-term subcutaneous EEG: a 15-month prospective study

**DOI:** 10.1101/2024.11.16.24317163

**Authors:** Pedro F. Viana, Jonas Duun-Henriksen, Andrea Biondi, Joel S. Winston, Dean R. Freestone, Andreas Schulze-Bonhage, Benjamin H. Brinkmann, Mark P. Richardson

**Author notes:** Correspondence: Dr. Pedro F. Viana, Institute of Psychiatry, Psychology & Neuroscience School of Neuroscience, Richardson Lab, King’s College London.

## Abstract

**Objective:** Novel subcutaneous electroencephalography (sqEEG) systems enable prolonged, near-continuous cerebral monitoring in real-world conditions. Nevertheless, the feasibility, acceptability and overall clinical utility of these systems remains unclear. We report on the longest observational study using ultra long-term sqEEG to date.

**Methods:** We conducted a 15-month prospective, observational study including ten adult people with treatment-resistant epilepsy. After device implantation, patients were asked to record sqEEG, to use an electronic seizure diary and to complete acceptability and usability questionnaires. sqEEG seizures were annotated visually, aided by automated detection. Seizure clustering was assessed via Fano Factor analysis and seizure periodicity at multiple timescales was investigated through circular statistics.

**Results:** Over a median duration of 438 days, ten patients recorded a median 18.8 hours/day, totalling 71,984 hours of real-world sqEEG data. Adherence and acceptability remained high throughout the study. While 754 sqEEG seizures were recorded across patients, over half (52%) of these were not reported in the patient diary. Of the 140 (27%) diary reports not associated with an identifiable sqEEG seizure, the majority (68%) were reported as seizures with preserved awareness. The sqEEG to diary F1 agreement score was highly variable, ranging from 0.06 to 0.97. Patient-specific patterns of seizure clustering and seizure periodicity were observed at multiple (circadian and multidien) timescales.

**Interpretation:** We demonstrate feasibility and high acceptability of ultra long-term (months-years) sqEEG monitoring. These systems help provide real-world, more objective seizure counting compared to patient diaries. It is possible to monitor individual temporal fluctuations of seizure occurrence, including seizure cycles.

**Summary for Social Media if Published:** [I do not have a X/Twitter handle]

People with epilepsy may suffer from severe injuries or sudden death due to uncontrolled seizures. Many seizures remain unnoticed by people and unreported to doctors, despite these potentially catastrophic consequences. There is an urgent need to detect seizures more objectively. Our study used a novel device placed under the skin to monitor brainwave activity continuously, at home, for many months, in people with treatment-resistant epilepsy. By monitoring epilepsy from home, we detected many seizures unreported by patients, and we also found predictable patterns (cycles) of seizure occurrence over time. In the future, this type of monitoring could revolutionize epilepsy care, by improving safety, treatment management and reducing uncertainty in patients’ daily lives.

## Introduction

In treatment-resistant epilepsy, accurate identification of seizures and other epilepsy-related objective disease information is a major challenge in clinical practice.^1^ Accumulated evidence demonstrates the unreliability of patient-reported seizure diaries.^1^ Patients may under-report a large proportion of seizures, while over-reporting some non-seizure symptoms as seizures. This can lead to inappropriate clinical management, from insufficient treatment exposing patients to seizure-related harms to excessive treatment contributing to unnecessary side-effects. Despite these limitations, seizure diary information remains the usual primary outcome measure for assessing efficacy of new treatments in clinical trials.^2^

Seizure unpredictability is also a major concern frequently highlighted in patient surveys.^3^ Although often perceived as completely random events, reports of non-random seizure timing date back millennia. Sir William Gowers classified epilepsy “chronotypes” (“nocturnal”, “diurnal” and “mixed”)^4^ and also described seizure clusters, or “groups of attacks”, suggesting that “seizures beget seizures”.^4^ Langdon-Down M. and others analysed in detail years-long seizure charts of patients in the Lingfield Epilepsy colony, finding not only characteristic circadian chronotypes but also periodic patterns lasting weeks, months or years.^5^

Tracking objective disease information and reducing seizure have motivated the development of mobile health monitoring systems.^6^ Their clinical applications are wide-ranging, from real-time seizure detection alarms^7^ to offline accurate seizure counting^8^ and seizure forecasting.^9–11^ Key requirements for the adoption of these systems include evidence that the technology is reliable, has suitable performance characteristics, addresses patient needs and is usable in the long-term.^12^

Available seizure detection devices mostly monitor indirect, non-cerebral biosignals as proxies for mostly major motor (tonic-clonic) seizures.^7,12^ In addition, despite multiple devices available on the market, evidence for their reliability and acceptability in the real-world is lacking.^13–15^

EEG remains the most important instrument in the evaluation of epilepsy, with the ability to characterise multiple seizure types, detect interictal epileptiform activity and monitor sleep.^16^ However, scalp EEG is limited to a few weeks at most, due to the potential for skin injury, inconvenience of visible electrode wires and signal quality degradation with time. Dry electrodes have been developed to overcome some of these limitations but signal quality also tends to degrade.^17^ Behind-the-ear or in-the-ear EEG have been studied in small numbers of patients but their long-term signal quality is unknown.^18,19^ Chronic invasive intracranial EEG systems have been developed as seizure warning systems^20^ or closed-loop stimulation devices,^21,22^ however they are associated with the risk of severe complications.

Subcutaneous EEG (sqEEG) could offer a trade-off between minimal invasiveness/low risk and good signal quality.^23,24^ sqEEG signal quality is similar to simultaneous scalp EEG^25^ and is highly stable over multiple months (ultra long-term).^24^ Recent single cases and case series have reported on the system’s feasibility, safety and spectrum of clinical indications.^8,26–29^

We report here on the longest prospective study (to the best of our knowledge) using ultra long-term sqEEG to date. We systematically assessed the system’s usability and acceptability, the diagnostic yield for seizures of different types, its comparison to patient diaries and utility to investigate individualised temporal dynamics of seizure occurrence.

## Methods

### Study Design and Population

The SUBER Study (“SUBcutaneous EEG: foRecasting of epileptic seizures through investigation of long-term dynamics of seizure occurrences, stress, sleep and other factors”) was an observational, prospective, non-randomized and non-interventional study, conducted at King’s College London and King’s College Hospital NHS Foundation Trust. Participants’ consent was obtained according to the Declaration of Helsinki and the study was approved by the local ethics committee (19/LO/0354). The patient IDs in this manuscript are not known to anyone outside of the research group.

### Patient Selection

We aimed to recruit ten adult people with treatment-resistant epilepsy (> 20 seizures per year according to seizure diary) of any syndrome, in which seizures were detectable in scalp EEG with two electrodes. We excluded patients with a diagnosis of potential seizure mimics (e.g. psychogenic non-epileptic seizures), significant medical comorbidities or with contraindication for placement of the subcutaneous EEG implant (e.g. planned MRI during the study period). The full inclusion/exclusion criteria can be found in **Supplementary Material 1**. Patients were recruited from epilepsy clinics at King’s College Hospital and St. George’s University Hospital. New participants were recruited if a participant dropped out prematurely from the study.

### Study procedures and data collection

A schematic of the main study procedures can be found in **Supplementary Material 2**. Patients pre-screened for eligibility criteria were approached during clinic or by a direct telephone call. Those interested in participating underwent an inclusion visit whereby eligibility criteria were confirmed, and pre-procedure laboratory blood tests performed.

Eligible patients were invited to a second visit during which the subcutaneous EEG device was implanted. Placement of the 24/7 SubQ implant involved a small 25mm incision made in the postauricular region using local anaesthesia and sterile technique. The electrode was placed in the subgaleal space, oriented towards the expected site of the strongest ictal EEG activity (determined after examining the participant’s previous investigations). More information about the implantation procedure can be found in Djurhuus BD et al.^30^

Data collection commenced one to two weeks after implantation. Patients were instructed to wirelessly connect the 24/7 EEG SubQ external logging device/data logger (**Supplementary Material 3**), which supplies the implant with power and stores recorded sqEEG. Patients were asked to connect the data logger for as long as possible during the day and night, except in circumstances when the device could get wet (e.g. personal hygiene). A 15-minute data quality protocol was conducted to record background activity, as well as common artefacts (e.g. eye blinks, chewing, eye movements, head movements, walking, running), to later facilitate the visual interpretation of the EEG (example recording in **Supplementary Material 4**).

Patients were asked to report their seizures using an electronic diary (Seer app, Seer Medical, Melbourne, Australia); they were given a Seizure Key card with description of each different seizure type. Diary entries were classified into three categories: non-convulsive with preserved awareness, non-convulsive with impaired awareness and convulsive/tonic-clonic. Participants were also requested to use a commercial-grade fitness tracker throughout the study (Fitbit^TM^ Charge 3 or 4, Fitbit, San Francisco, USA) that estimates heart rate, step counts, sleep duration and sleep staging.

Up-to-monthly follow-up visits were conducted to offload sqEEG data, review patient safety, experiences with the device, seizure diaries and treatment changes. Patients completed the brief Illness Perception Questionnaire^31^ at baseline (**Supplementary Material 5**). Participant satisfaction with the sqEEG system was measured at 3 months post-implantation and at study end. Overall satisfaction was assessed via a 7-item Likert scale questionnaire (Fig 1C). Usability was assessed with a 10-item System Usability Scale (SUS, Fig 1D).^32^ Clinical care was not altered by participation in the study, hence medication and other treatment changes were allowed. Patients were encouraged to collect data but no specific study adherence criteria for patient drop-out were set. Device explantation at end of the study was performed under local anaesthesia during a half-day hospital visit. Adverse events and device deficiencies were collected throughout the study.

**Figure 1.**
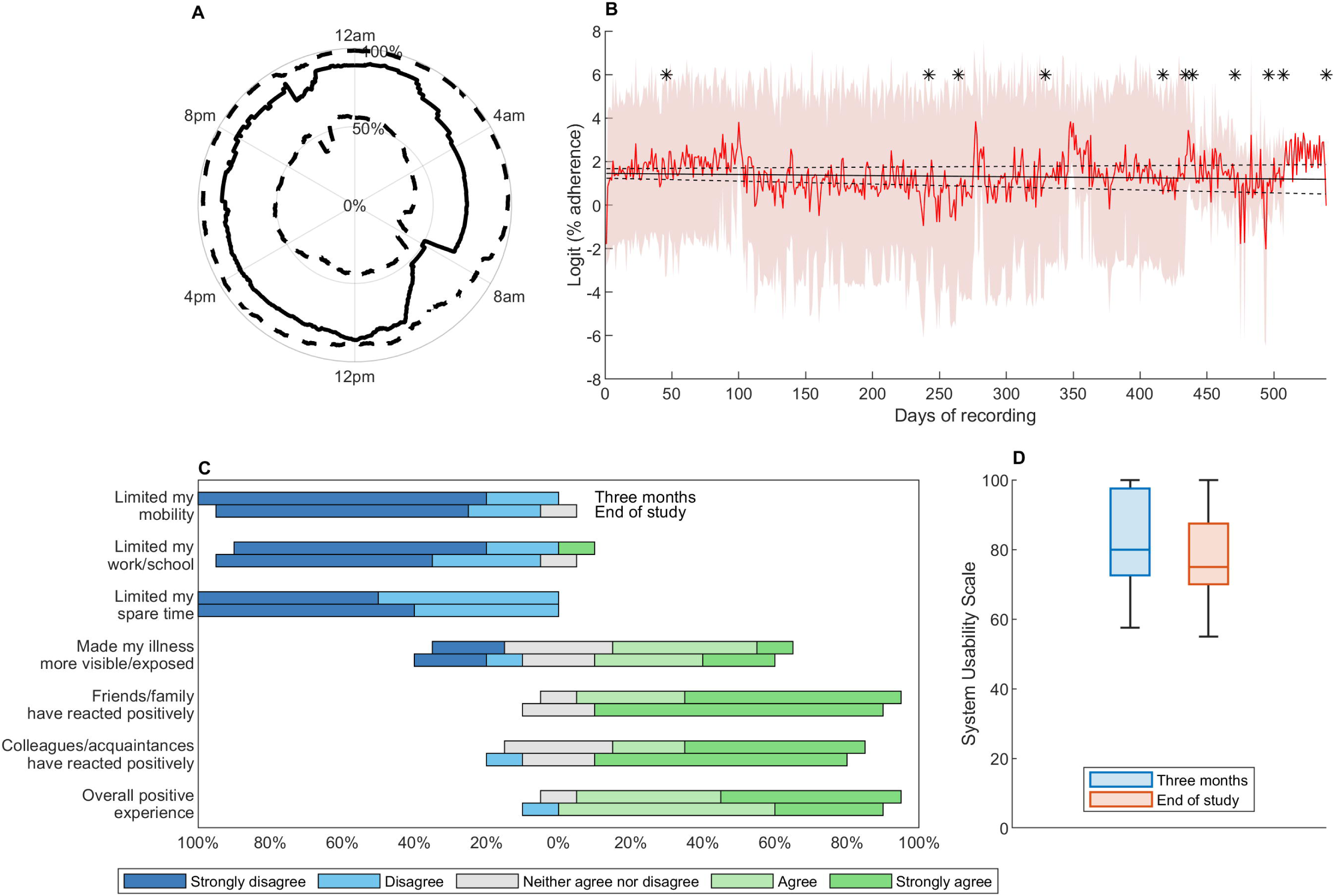
Adherence and acceptability of ultra long-term sqEEG in our cohort. (**A**) Circadian polar plot showing median (solid line) and interquartile ranges (dashed lines) of average adherence at different times of day, across study participants. (B) Long-term adherence. Red line and shaded area represent the mean and standard deviation of logit daily adherence, across participants. Black asterisks denote the end of the recording for each subject. The black solid line represents the result of the group-level linear model, and dashed lines are 95% upper and lower bounds of the model. **(C)** Results of the Acceptability Questionnaires at three months and at study end. **(D)** System Usability Scale at three months and study end.

The study procedures underwent several modifications throughout the study period:

– A malfunction of the implant was encountered, compromising data quality and preventing further recording in the first two patients of the study (after 9 months and 1.5 months of recording). The study was paused until the device company manufactured and had CE-approval of a more robust implant. One patient agreed to be re-implanted and the other dropped out of the study.
– The UK government-mandated lockdown due to the Covid-19 pandemic resulted in a longer interruption. The study reopened with several modifications, including 1) possibility of remote (telephone) follow-up visits and 2) remote sqEEG data collection, whereby the data was uploaded by the patient onto a USB-drive, using a customized laptop and software (UNEEG Home Data Manager System), and the USB-drive containing anonymised data was shipped regularly via post.
– To compensate for study interruptions and missing data, participants were given the option to extend monitoring, from 12 months as initially planned to 15 months prior to device explantation.

### Data pre-processing and seizure annotations

The two-channel subcutaneous EEG signal is recorded at a sampling rate of 207Hz and bandpass filtered at 0.5 - 48Hz with a finite-impulse-response equiripple design and 40 dB attenuation filter, prior to review. The sqEEG was reviewed on a dedicated software (UNEEG Episight viewer – example in **Supplementary Material 7**) with an in-built 10-minute spectrogram viewer and a high-sensitivity data review reduction seizure detector. Two seizure detection algorithms were used in the study: 1) an initial version (v1.11) used in the first participant and 2) an improved and published version (v2.0) used in the remaining participant data.^33^ Electrographic seizures were identified by a board-certified electroencephalographer (P.F.V.) and an experienced EEG technologist (Christian Skaarup, UNEEG Medical A/S). Several examples per patient were discussed and reviewed with a second board-certified clinical neurophysiologist (J.S.W.). Patient-specific electrographic seizure patterns (seizure signatures), taken from previous recordings, were initially assessed by reviewers to facilitate their identification on sqEEG. The sqEEG quality protocol recordings were also useful at identifying potential seizure mimics due to common rhythmic artefacts (e.g. movement artefact during walking). Electrographic seizures were further classified into convulsive and non-convulsive, as previous preliminary work showed perfect inter-rater agreement to differentiate between both seizure types (**Supplementary Material 8**).

The sqEEG data review process was conducted as follows: 1) review of events marked by the high sensitivity seizure detector; 2) review of periods around the patient diary reports (within two hours pre and post report), and 3) review of a random sample of 6-hour epochs (in 10-minute spectrogram epochs) comprising 10% of the whole recording. Finally, the full dataset (in 10-minute spectrogram epochs) was reviewed if sensitivity of the seizure detection algorithm was found to be below 80%.

### Statistical Analyses

Descriptive analysis was summarized using median and interquartile range for continuous variables and absolute counts (percentages) for categorical variables. Adherence to sqEEG was assessed at circadian, weekly and long-term timescales. A day-of-the-week effect was investigated at individual level via Kruskal-Wallis tests. To assess long-term adherence to sqEEG, linear regression models were constructed, both at group level and individual levels. The outcome variable was percentage adherence per calendar day (after logit transformation), and the dependent variable was time (in days since start of recording).

Comparison to diary records was made by matching every sqEEG seizure with the closest diary event, if the event was reported in the vicinity (i.e. +/-two hours) of the seizure. The F1 score was calculated to assess the overall agreement between diary events and sqEEG seizures.

Seizure clustering was assessed via inspection of cumulative seizure count plots, as well as by calculating the Fano Factor (ratio of the variance to the mean of seizure frequency) across daily, weekly and monthly timescales, for both the diary and sqEEG.^34,35^ For a Poisson process, the Fano Factor = 1; a clustering process exhibits Fano Factor > 1, and for a regular periodic process it is < 1. Significance was calculated using a previously described method based on the gamma distribution.^34,36^ Clustered seizures were defined as those preceded by at least one seizure in the last 24 hours (see varying seizure cluster definitions in ^37^).

We analysed the temporal periodicity of both sqEEG seizures and diary events, adapting from previous work on seizure cycles.^38,39^ We included patients with >10 seizures and/or diary events, and with more than 50% of adherence throughout the study, to minimize observing spurious periodicity due to non-random low device adherence. Seizure periodicities were analysed over multiple timescales (from 6h to 48h at 6h intervals, from 3 days up to 20% of the recording duration at 1-day intervals).^38,39^ The synchronisation index, the sum of unit vectors with angle representing each seizure in the cycle, was calculated for each cycle.^38^ Statistical significance was determined by the Omnibus test for non-uniformity of angles in a circle, after multiple comparisons correction by the Benjamini-Hochberg False Discovery Rate procedure.^40^ Significance was assessed at q-value<0.05. Data analysis was performed using Microsoft Excel 365 and MATLAB (MathWorks, R2024a, USA).

## Results

### Study Population

Amongst fifteen patients who initially consented to participate in the study, two (SK, SO) failed screening and one (SL) withdrew consent before the implantation visit. Of the twelve patients implanted, two dropped out before recording usable data – one whose implant was misplaced during surgery, and with skin protrusion necessitating removal, and who did not wish to be reimplanted (SM), and one who reported an immediate headache and scalp pain post-implantation who did not wish to wait for improvement (SN). As previously mentioned, a device malfunction early in the study led to an early drop-out of one participant (SB, recording 45 days), while another patient agreed to be reimplanted (SA -> SA2). Hence, the final cohort with usable data included 11 datasets from 10 patients (Table 1). All patients had focal epilepsy of structural or presumed structural (i.e. MRI-negative) aetiology. Age ranged from 29 to 64 years, and half were male. All were on at least two regular anti-seizure medications, and two participants also had active vagus nerve stimulation (SF, SI). The majority (7/11) of implant locations were left-sided.

**Table 1.**
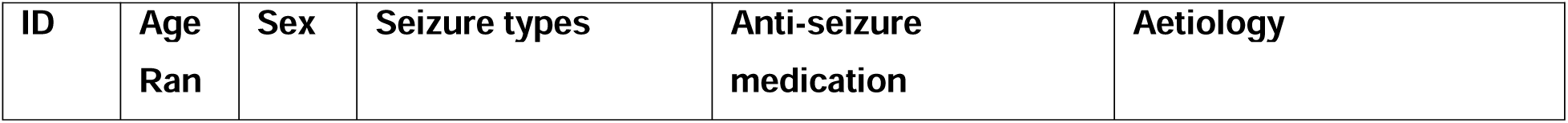
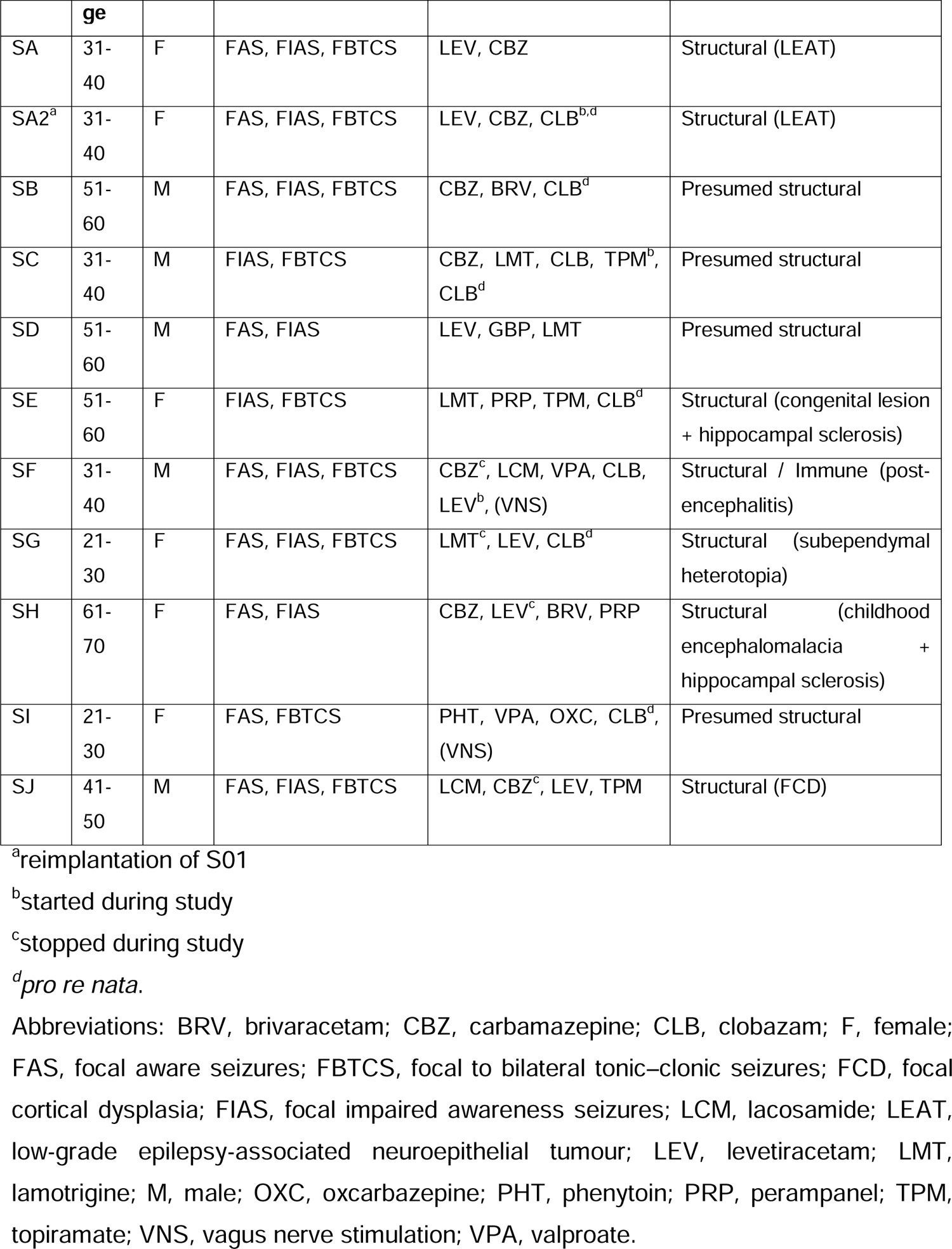
Demographic and clinical characteristics of the cohort.

| ID | Age<br>Ran | Sex | Seizure types | Anti-seizure<br>medication | Aetiology |
| --- | --- | --- | --- | --- | --- |

|  | ge |  |  |  |  |
| --- | --- | --- | --- | --- | --- |
| SA | 31-40 | F | FAS, FIAS, FBTCS | LEV, CBZ | Structural (LEAT) |
| SA2<sup>a</sup> | 31-40 | F | FAS, FIAS, FBTCS | LEV, CBZ, CLB<sup>b,d</sup> | Structural (LEAT) |
| SB | 51-60 | M | FAS, FIAS, FBTCS | CBZ, BRV, CLB<sup>d</sup> | Presumed structural |
| SC | 31-40 | M | FIAS, FBTCS | CBZ, LMT, CLB, TPM<sup>b</sup>, CLB<sup>d</sup> | Presumed structural |
| SD | 51-60 | M | FAS, FIAS | LEV, GBP, LMT | Presumed structural |
| SE | 51-60 | F | FIAS, FBTCS | LMT, PRP, TPM, CLB<sup>d</sup> | Structural (congenital lesion + hippocampal sclerosis) |
| SF | 31-40 | M | FAS, FIAS, FBTCS | CBZ<sup>c</sup>, LCM, VPA, CLB, LEV<sup>b</sup>, (VNS) | Structural / Immune (post-encephalitis) |
| SG | 21-30 | F | FAS, FIAS, FBTCS | LMT<sup>c</sup>, LEV, CLB<sup>d</sup> | Structural (subependymal heterotopia) |
| SH | 61-70 | F | FAS, FIAS | CBZ, LEV<sup>c</sup>, BRV, PRP | Structural (childhood encephalomalacia + hippocampal sclerosis) |
| SI | 21-30 | F | FAS, FBTCS | PHT, VPA, OXC, CLB<sup>d</sup>, (VNS) | Presumed structural |
| SJ | 41-50 | M | FAS, FIAS, FBTCS | LCM, CBZ<sup>c</sup>, LEV, TPM | Structural (FCD) |

<sup>a</sup>reimplantation of S01
<sup>b</sup>started during study
<sup>c</sup>stopped during study
<sup>d</sup>*pro re nata*.
Abbreviations: BRV, brivaracetam; CBZ, carbamazepine; CLB, clobazam; F, female; FAS, focal aware seizures; FBTCS, focal to bilateral tonic-clonic seizures; FCD, focal cortical dysplasia; FIAS, focal impaired awareness seizures; LCM, lacosamide; LEAT, low-grade epilepsy-associated neuroepithelial tumour; LEV, levetiracetam; LMT, lamotrigine; M, male; OXC, oxcarbazepine; PHT, phenytoin; PRP, perampanel; TPM, topiramate; VNS, vagus nerve stimulation; VPA, valproate.

### Device Safety and Device Deficiencies

Twelve adverse events (AEs) occurred throughout the study, of which seven were deemed possibly or probably related to study participation (**Supplementary Material 9**). Most adverse events were mild and comprised of temporary pain/headache after the implantation procedure. Two serious AEs occurred throughout the study – one hospitalization due to community acquired pneumonia (unlikely related to study participation) and one unanticipated hospitalization to urgently remove a misplaced implant causing tip protrusion through the scalp (hospitalization was only necessary due to Covid-19 restriction measures). All AEs were associated with recovery without any sequelae.

Thirty-two device deficiencies (**Supplementary Material 10**) occurred throughout the duration of the study, of which 25 affected sqEEG data collection. Most deficiencies were related to the external data logger, only temporarily affected data collection and were resolved by rebooting or (more rarely) replacement of the devices.

### Device Adherence and Acceptability

For a median recording duration of 433 days, participants recorded a median of 18.8 (IQR 12.1) hours per day (i.e. 78.4% of the recording time – Table 2). Five participants recorded over 20h/day, while three recorded under 12h/day. One participant (SI) had simultaneous implantation of a VNS device and experienced unanticipated freedom from impaired awareness seizures, which limited his motivation to record. Overall, 71,984 hours of real-world sqEEG were collected.

**Table 2.** Implantation and recording characteristics of the cohort.

| **Subject ID** | **Implant Location** | **N. hours of sqEEG recorded** | **Recording duration (days)** | **Data capture rate (%)** | **Data capture rate (hours/day)** | **N. seizures recorded** |
| --- | --- | --- | --- | --- | --- | --- |
| SA | LT | 4941 | 241 | 85.4% | 20.5 | 33 |
| SA2<sup>a</sup> | LFT | 9319 | 495 | 78.4% | 18.8 | 92 |
| SB | RT | 809 | 45 | 74.7% | 17.9 | 36 |
| SC | RFT | 6943 | 506 | 57.2% | 13.7 | 56 |
| SD | RFT | 10682 | 470 | 94.7% | 22.7 | 54 |
| SE | LFT | 11692 | 538 | 90.5% | 21.7 | 54 |
| SF | LFT | 10363 | 438 | 98.6% | 23.7 | 132 |
| SG | LFT | 2783 | 263 | 44.1% | 10.6 | 203 |
| SH | LFT | 3620 | 433 | 34.8% | 8.4 | 31 |
| SI | LC | 1360 | 329 | 17.1% | 4.1 | 0 |
| SJ | RFT | 9472 | 417 | 95.0% | 22.8 | 63 |
| **Total** | **-** | **71984** | **-** | **-** | **-** | **754** |
| **Median (IQR)** | **-** | **2783 (6943-10363)** | **433 (263-495)** | **78.4% (44.1-94.7%)** | **18.8 (12.1)** | **54 (33-92)** |

<sup>a</sup>reimplantation of S01
Abbreviations: LT – left temporal; LFT – left frontotemporal; RT – right temporal; RFT – right frontotemporal; LC – left central

Circadian adherence plots (**Supplementary Material 11**) showed highly individualised patterns of adherence, from strictly fixed hours off recording in some participants, to a preference to record either during the day or night in others. A weekday effect was statistically significant in four participants (**Supplementary Material 12**). Analysing long-term adherence, a group-level model showed no significant attrition throughout the study (Fig 1B), and most individual long-term adherence models also showed a temporal trend close to zero **(Supplementary Material 3).**

Acceptability and usability of the system was overall high and remained high throughout the study (Figs 1C-D). In general, participants felt that the system did not limit their daily lives. About half of participants felt that the system made their chronic illness more exposed, however the general impression from family, friends or colleagues was positive.

### Recorded Seizures and Comparison to Diary

Example recordings comparing sqEEG and diary events can be found in Fig 2. After final data review, 754 seizures were annotated in total across participants (Table 2). The number of seizures per participant ranged from zero (SI with low adherence and reported freedom from impaired awareness seizures) to 203. A small number of convulsive seizures (n=10) was recorded in three participants. Each patient (and seizure type) had their own ictal seizure signature visible on the EEG time series and spectrogram (**Supplementary Material 14**).

**Figure 2.**
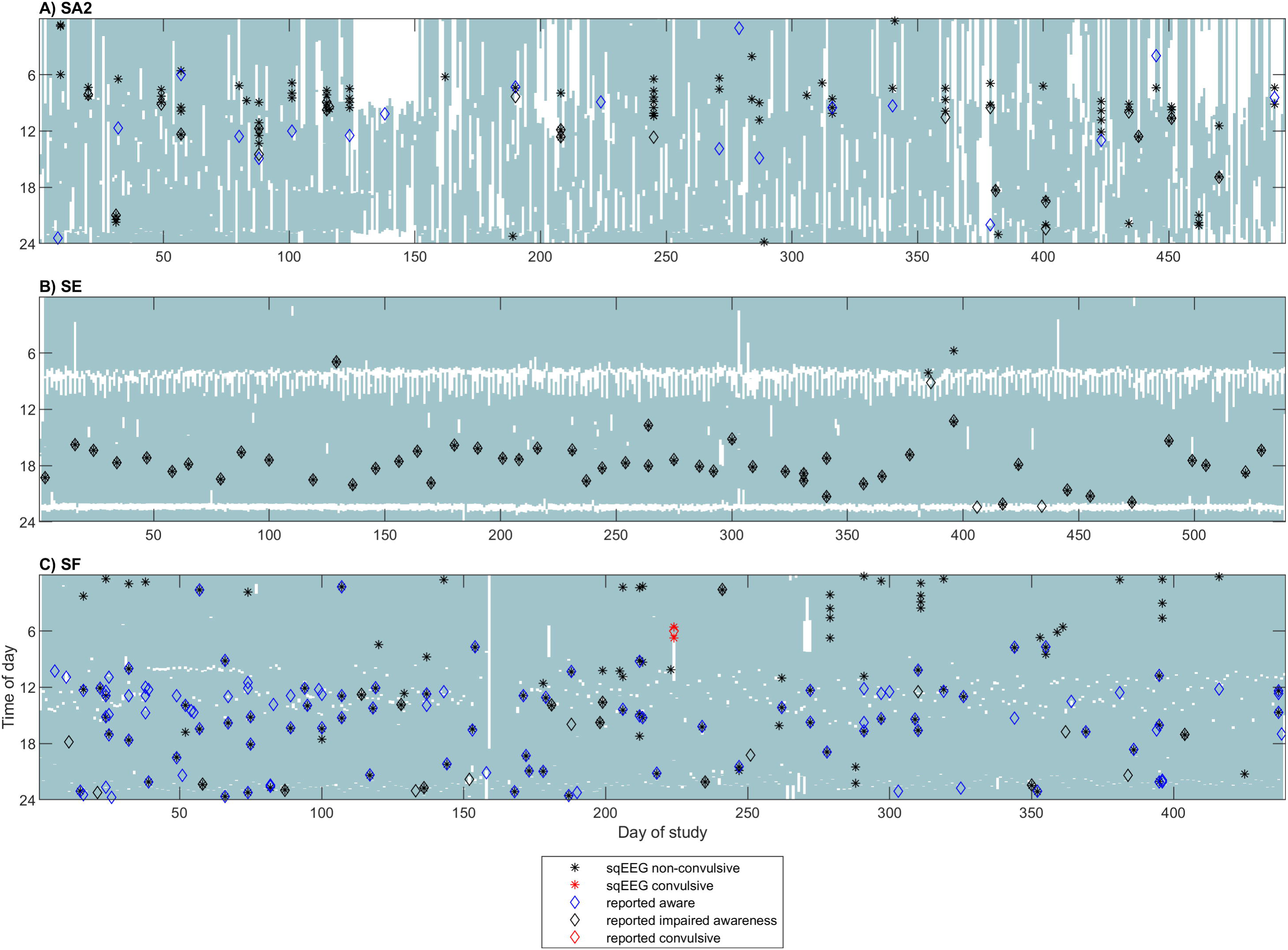
Example participant recording day of study (x-axis) vs. time of day (y-axis) plots. The shaded areas indicate periods when the patients were recording. Diamonds represent times of diary reported aware (blue), impaired awareness (black) and convulsive (red) seizures. Asterisks indicate times of non-convulsive (black) and convulsive (red) seizures. (**A**) SA2, showing examples of clustered seizures (vertical columns of multiple asterisks), as well as a mixture of well-reported with missed sqEEG seizures. Note the circadian (morning) preference for seizure occurrence. (**B**) SE, as an example of a very strict device adherence pattern, perfect diary-to-sqEEG agreement, absence of seizure clustering and circadian (early evening) preference for seizure occurrence. **(C)** SF, showing many reported (mostly aware) events without a sqEEG correlate; many non-reported nocturnal sqEEG seizures; one non-reported clustered tonic-clonic sqEEG seizure.

Conversely, 592 events were reported in participants’ seizure diaries, of which 506 were reported at times when the sqEEG device was being used. Most diary reports were reported as non-convulsive impaired awareness (n=237) and non-convulsive aware seizures (n=236).

After matching individual sqEEG seizures with the closest diary events (within two hours from each other), we found that over half (52%) of recorded sqEEG seizures were not reported in the diary, including three convulsive seizures (Table 3). Conversely, 140 diary events were not associated with a sqEEG seizure. Most (68%) non-recorded diary events were reported as aware seizures, and no reported convulsive seizures were missed by the sqEEG device. In addition, there were discrepancies between the type of seizure reported in the diary and the pattern detected on sqEEG. Across patients, the F1 agreement score between reporting a diary event and recording a sqEEG seizure was 0.58, whilst the within-patients median F1 score was 0.56, ranging from 0.06 (SI) to 0.98 (SE).

**Table 3.** Comparison between diary reported events and sqEEG recorded seizures.

|  |  | sqEEG |  |  | Total |
| --- | --- | --- | --- | --- | --- |
|  |  | Nonconvulsive | Convulsive | Not recorded |  |
| **Diary** | Aware | 141 (19%) | 0 (0%) | 95 (68%) | **236** |
|  | Impaired awareness | 201 (27%) | 1 (10%) | 35 (25%) | **237** |
|  | Convulsive | 3 (0.4%) | 6 (60%) | 0 (0%) | **9** |
|  | Unclassified | 14 (1.8%) | 0 | 10 (7%) | **24** |
|  | Unreported | 385 (52%) | 3 (30%) | - | **388** |
| **Total** |  | **744** | **10** | **140** |  |

### Patterns of Seizure Occurrence

The proportion of clustered seizures (seizures occurring within 24h of a preceding seizure) ranged from 2% (SD) to 87% (SG), with a median of 42%. In addition, a subset of participants had increased Fano Factor values consistent with seizure clustering at different timescales (Fig 3A). Conversely, some participants (e.g. SE) had Fano Factor values <1, in keeping with a periodic pattern of seizure occurrence. Fano factors were more frequently significant when analysing the sqEEG, compared to the diary (Fig 3A). Analysing seizure periodicities (Fig 4), most participants included in the analysis had significant circadian cycling of seizure occurrence, either evident from the seizure diary or sqEEG. In addition, four participants showed significant multidien cycles. In several participants, there was a marked similarity between diary and sqEEG seizure cycling. However, some discrepancies were found. For example, in SF the diary suggested a strong 24h cycle, whilst the sqEEG data showed no significant periodicity. Analysing the participant’s recording (Fig 2C), it was evident that the patient was not reporting many nocturnal seizures, which likely led to an overestimation of a diurnal preference for seizure occurrence.

**Figure 3.**
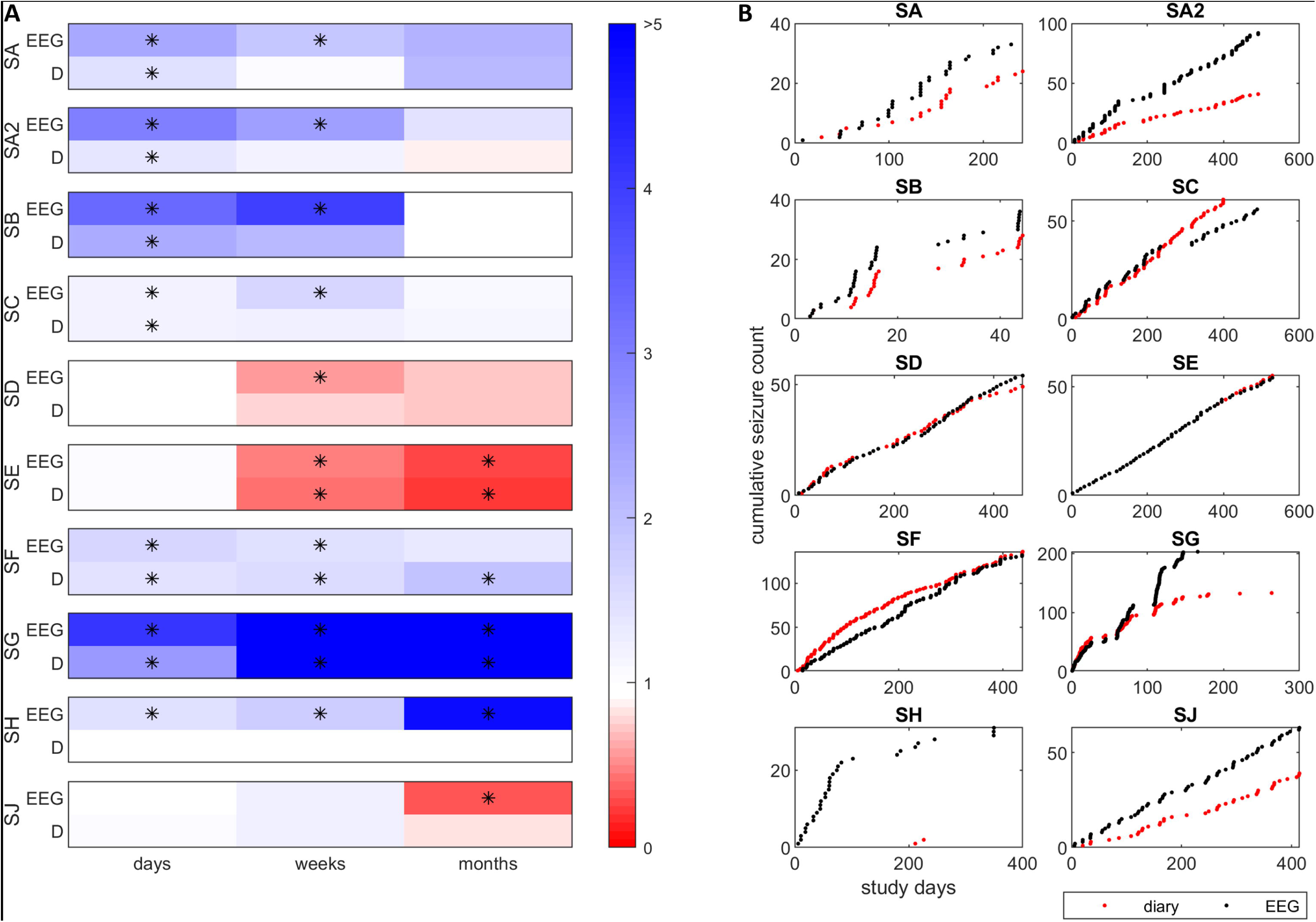
Seizure clustering in the SUBER cohort. **(A)** Fano Factor (FF) values for each participant at day, week and month timescales, based on the sqEEG and diary (“D”) data. A Fano Factor of 1 corresponds to an expected Poisson distribution. Asterisks indicate significant (p<0.05) deviation from FF=1. Note, for example, FF values higher than 1 in SA, SB and SA2, suggesting a high degree of clustering, and FF lower than 1 in e.g. SE suggesting a more periodic non-clustering pattern. **(B)** Scatter plots for the different participants showing seizure occurrence vs. cumulative seizure counts, for diary and sqEEG data, showing clustering patterns in some (SA, SA2, SB, SG) participants compared to others without clustering (SD, SE, SF).

**Figure 4.**
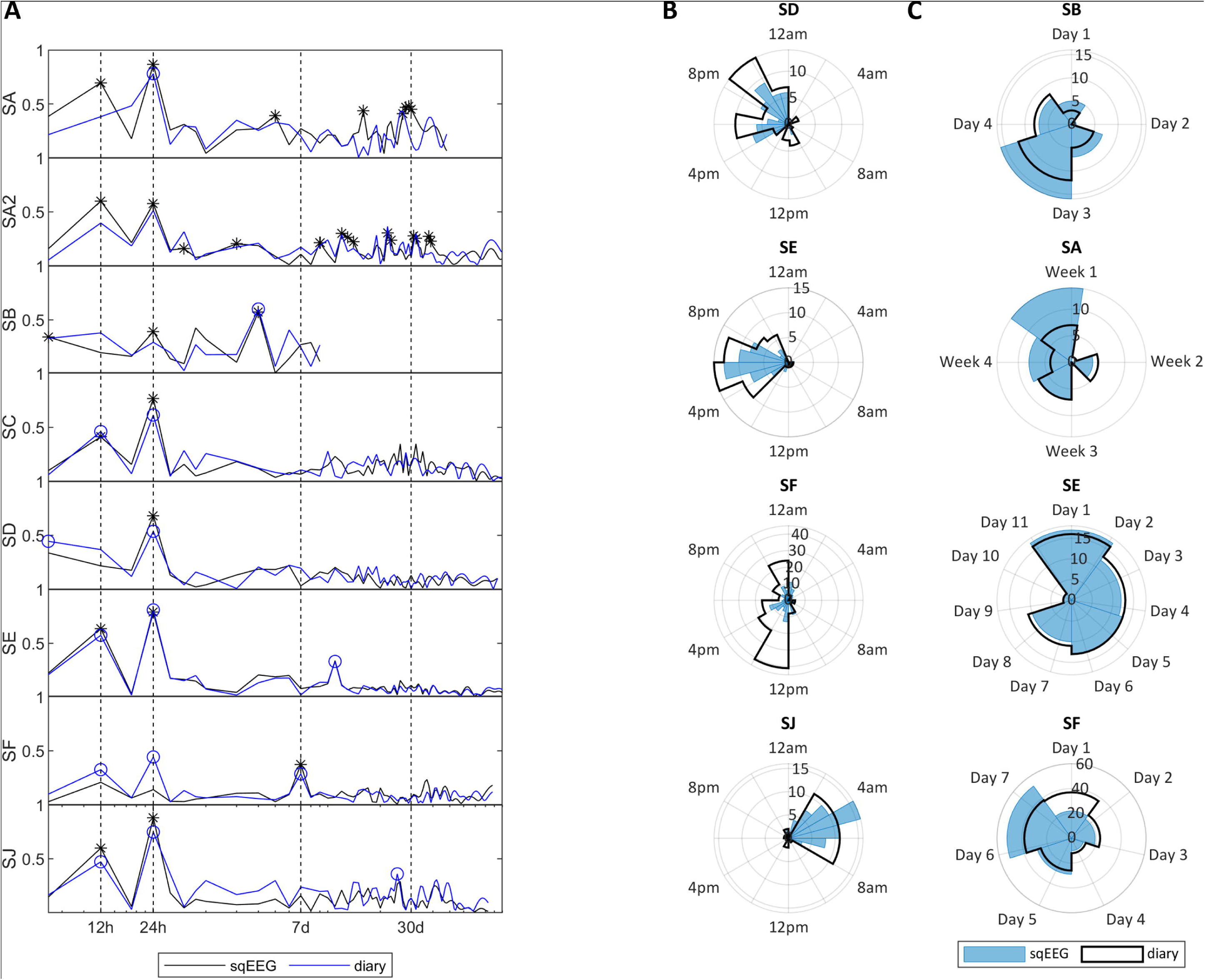
Seizure cycles in the SUBER cohort. **(A)** Distribution of the synchronization index of sqEEG seizures (black lines, significant cycles with asterisks) and diary reports (blue lines, significant cycles with circles), for different participants, at different cycle lengths. Note the logarithmic scale in the x-axis. **(B)** Example polar histograms of sqEEG seizures (blue) and diary events (black outline) distributed over circadian (24h) and **(C)** multidien cycles. Concentric rings represent number of dairy events or seizures.

### Performance of automated seizure detector

Table 4 shows the performance of the employed seizure detection algorithm aimed at reducing the amount of data needing manual review. Performance was quite variable between participants, with a median (IQR) sensitivity of 70.5% (48-94.6%) and pre-review false detection rate of 3.4/day (3-15.8/day). Performance was excellent in a group of patients (e.g. SC, SE, SJ) but poor in others (e.g. SB, SD, SF) which required full review of their datasets (see also **Supplementary Material 15**).

**Table 4.**
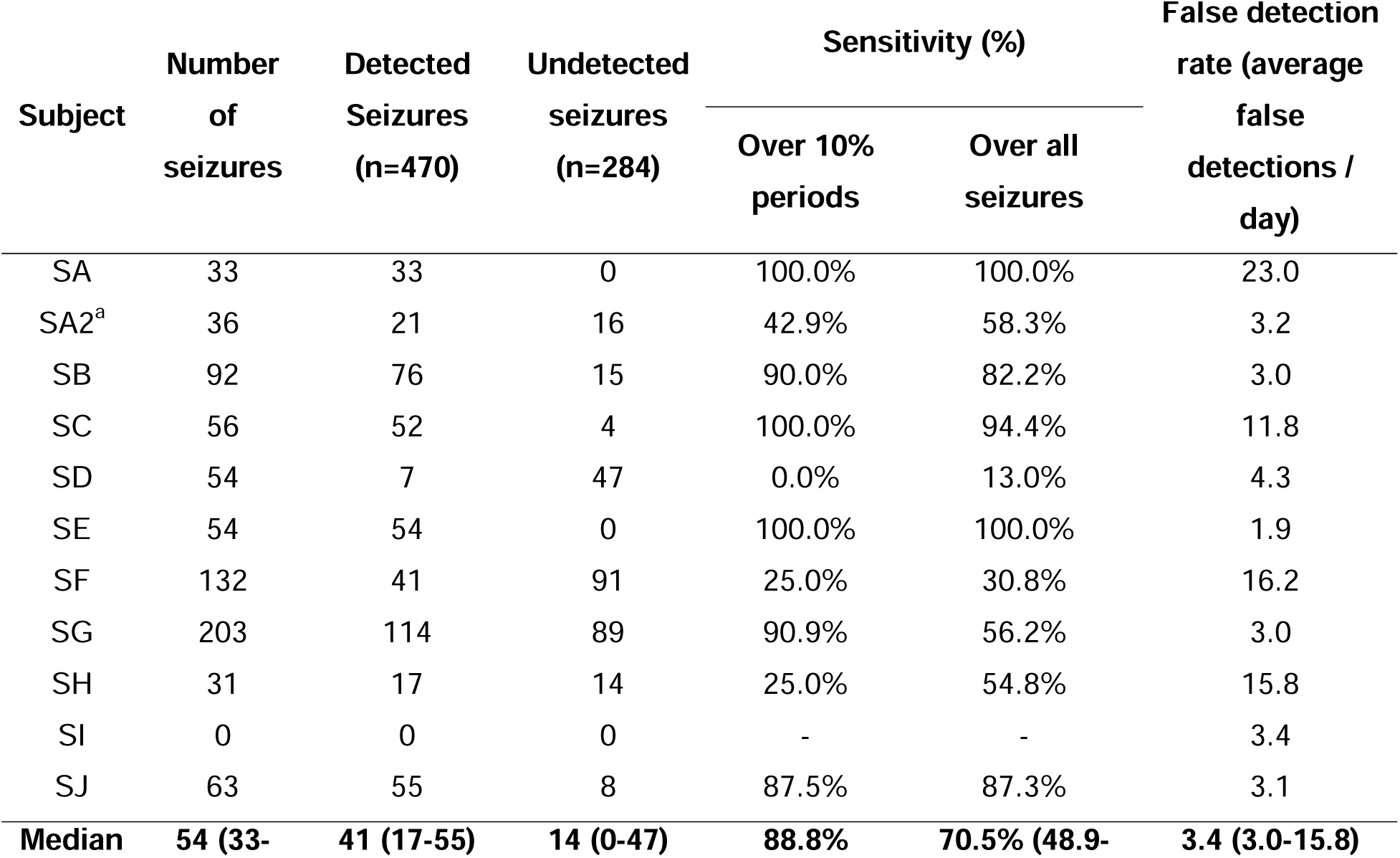
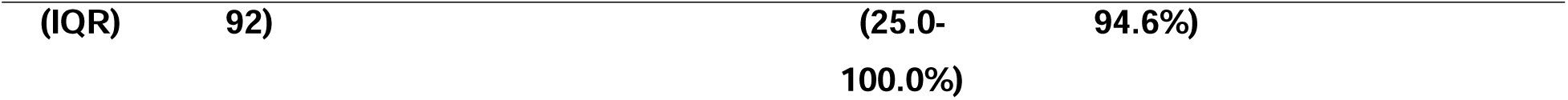
Performance of the data-reduction seizure detection algorithm.

## Discussion

To the best of our knowledge, we conducted the longest systematic prospective observational study using ultra long-term sqEEG to date, recording over 70,000 hours of EEG from 10 patients, spanning up to 15 months each, and including over 700 confirmed seizures. We demonstrated the feasibility of ultra long-term sqEEG monitoring, and we highlight important aspects of clinical applicability of these systems, namely the improvement in documentation of different seizure types, aided by automated seizure detection, and insights into personalised long-term temporal dynamics of seizure occurrence.

The implantation of the sqEEG device was well tolerated by most patients in the study. Most side-effects were mild, transient and related to the local anaesthetic procedure. However, there was one significant complication after surgery (electrode erosion through the skin) and one patient dropped out early due to localized headache. Early malfunction of the sqEEG device led to an additional study drop-out. These are all important learning points for both future researchers and device manufacturers. Progressive training on the surgical procedure and manufacture of more robust devices will probably mitigate some of these adverse events in the future.

Our recordings had overall high adherence, higher than reported in a previous three-month trial.^29^ At the group level we did not see significant attrition rate in recording adherence, contrary to what is often reported in device studies.^41^ Half of our cohort had excellent adherence (>20 hours/day). This shows that at least a proportion of patients is highly motivated to use these devices during their daily lives. Nevertheless, there were patients with poor adherence to the device. This might reflect the inconvenience and possible stigma of wearing a data logger, or the lack of perceived direct clinical benefit during this observational study. These points highlight the need to develop and improve devices that are less obtrusive, less stigmatizing and, above all, that translate to a direct clinical benefit to the patient.^12^ Participants also showed individual patterns of adherence, both at circadian and weekly levels. This unbalanced adherence is important to consider, particularly when interpreting temporal trends of both seizures and interictal activity.

When comparing sqEEG seizures with diary events, expected discrepancies were encountered. Patients failed to report over half of recorded seizures. A third of tonic-clonic sqEEG seizures were not associated with a diary report. This level of under-reporting is consistent with previous studies comparing seizure diaries against video-EEG^1^ and ambulatory EEG.^20,42^ Reasons for under-reporting have not been thoroughly investigated. Patients may lack the perception to report a seizure (e.g. seizure with impaired awareness and no warning or subjective symptoms), may forget a seizure occurred (e.g. seizure with retrograde amnesia) or may be unable to document it (e.g. due to temporary cognitive or motor impairment). Patients with particularly high seizure frequency may lose interest and motivation to track all seizures.

Conversely, 28% of diary reports were not associated with a recorded sqEEG seizure. The two-channel unilateral device may have limited spatial sampling to detect some distant and/or contralateral seizures. Most non-recorded diary events were reported as aware seizures, for which even full-scalp EEG has limited sensitivity.^43^ Some seizures recorded in the real-world may be obscured by activities of daily living and associated movement/muscle artefacts (examples in Viana PF et al^28^). Importantly, over-reporting is an often overlooked but clinically relevant aspect in diary seizure misdocumentation. Reasons may include the reporting of psychogenic non-epileptic seizures or of other paroxysmal events unrelated to seizures (e.g. cardiogenic syncope, panic attacks).

In this study, an automated seizure detection algorithm was used to reduce the amount of data to be reviewed to facilitate and accelerate the seizure annotation process. Across participants, the algorithm’s sensitivity was well within the reported range for most scalp EEG-based seizure detection algorithms (median 88.8% against a reported range of 75-90%)^44^, with also an acceptable pre-review false detection rate (median 3.4/day against a reported range of 2.4-120/day).^44^ The algorithm had excellent sensitivity in a proportion of participants, enabling significant reduction of the amount of data reviewed manually. However, sensitivity was low in a few patients for whom comprehensive manual review was eventually needed (Table 4 and **Supplementary Material 15**). These examples highlight a clear need for the improvement of automated seizure detection methods, particularly applied to limited channel real-world EEG data. Future directions may include the incorporation of patient-specific/personalized seizure patterns, well described in scalp^45^ and subcutaneous EEG data.^28,46^

One advantage of using sqEEG for seizure counting is to characterise different seizure types within the same patient, allowing phenotyping and risk stratification. Seizures with tonic-clonic artefacts were easily distinguishable from non-convulsive seizures (**Supplementary Material 8**), and good discrimination based on singe channel surface EMG electrodes has been previously described.^47^ Further more refined characterisation of seizure types, based on patient-specific artefact patterns^28^ or seizure duration^48^ may be possible with two-channel data and should be the subject of future work.

Chronic EEG recordings have enabled the characterization of temporal dynamics of seizure occurrence at the individual level.^34,35,39^ Similarly to previous reports, we have observed individualised patterns of both seizure clustering and periodicity. The proportion of clustered seizures compared to lead seizures varied widely between patients, from 2-87%, and the degree of clustering at different timescales showed high inter-individual variability, but apparently high intra-individual stability (Fig 3). In addition, many seizure clusters were missed by seizure diaries, where often only one seizure was reported (examples in Fig 2). Identifying seizure clusters may have additional utility to prompt administration of short-acting medication, particularly if the detection is timely.^37^

We have shown that it is also possible to determine individual seizure periodicities using sqEEG. Circadian periodicity was very common in our cohort, and significant multidien cycles were identified in half of the patients, ranging from timescales of a few days to approximately monthly. It is interesting to note that, even though over half of sqEEG seizures weren’t reported in the diary, in many patients the shape of the distribution of seizure cycles between both modalities was similar. Nevertheless, several periodicities were only significant on sqEEG, whilst some were identified from diary data. These discrepancies may reflect biased misreporting from the diary (example in Figure 2C) while, on the other hand, may be related to (biased) discontinuities in the recordings. Forecasting performance has been shown to improve when periodicity is included in prediction models.^11^ Future studies should further explore the feasibility and method of incorporating sqEEG seizure cycles into prospective forecasting. One option would include displaying polar plots in sqEEG reports (examples in Fig 4) for patients to be aware and for clinicians to consider adjusting medication timing. Another option includes incorporating daily (or even hourly where appropriate) seizure risk into a user’s daily life calendar.^49^

This study has other limitations in addition to those reported above. The sample size is small and is not necessarily representative of the population of people with treatment-resistant epilepsy. The placement of the subcutaneous EEG implant was determined after examining the patients’ previous video-EEG investigations, but no period of simultaneous video-EEG and sqEEG was performed. We found it unfeasible to perform this in our study due to clinical demand for prolonged video-EEG in our centre and the likely need for medication reduction in some patients to capture seizures within a conventional video-EEG study timeframe (e.g. the mean seizure frequency in SH was 2/month). Anti-seizure medication purely for research reasons would be ethically challenging given the associated risks.

A significant challenge will be to determine how to both implement and deliver this service within a healthcare system. Review of event detections is time consuming and requires specific expertise. After review, there will need to be a feasible way to relay the information back to both the patient and clinician. This could take the form of a periodic report, e.g. comparing a recent period with a reference. How frequently the information should be updated and shared remains to be determined.

In conclusion, we demonstrated that ultra long-term subcutaneous EEG recordings are feasible and may provide a range of clinically useful information to patients with drug-resistant epilepsy and their clinicians. It is possible to detect seizures more objectively, during routine daily life out of hospital. Recorded seizures can be categorized into different subtypes which, together with other EEG biomarkers, could allow more objective and timely disease severity stratification. It is possible to monitor individual temporal fluctuations of seizure occurrence, including seizure cycles. Overall, these findings, conducted in a small group of patients but over a long time period, show the potential of sqEEG for a diverse range of clinical applications, from automated seizure detection to seizure prediction. This work calls for future, larger scale, prospective trials to further validate this technology.

## Data Availability

Data from selected subcutaneous EEG recordings that support these findings will be made available in a future open-source seizure prediction challenge.

## Acknowledgements

This work was supported by the Epilepsy Foundation’s Epilepsy Innovation Institute My Seizure Gauge Project. MPR is supported by the NIHR Biomedical Research Centre; the MRC Centre for Neurodevelopmental Disorders (MR/ N026063/1). BHB was supported by the NIH NINDS (UG3 NS123066). We would like to thank the Neurosurgical team (Mr. Harishchandra Srinivasan, Mr. Harutomo Hasegawa and Mr. Richard Selway) involved in the implantation procedures at King’s College Hospital NHS Foundation Trust. We would like to acknowledge Christian Skaarup and Cecilie Bottiger (UNEEG Medical A/S) for their valuable technical support. Ultimately, we would like to acknowledge all patients who participated in this study.

## Author Contributions

Conception and design of the study: PFV, JDH, DRF, ASB, BHB, MPR. Acquisition and analysis of data: PFV, JDH, AB, JSW. Manuscript draft: PFV.

## Abbreviations

IQR: interquartile range

KCL: King’s College London

PWE: people with epilepsy

sqEEG: subcutaneous EEG

SUS: system usability scale

VNS: vagus nerve stimulation.

