## Supplementary Data for "Real-world epilepsy monitoring with ultra long-term subcutaneous EEG: a 15-month prospective study"

Supplementary Material

### 1. Study Eligibility Criteria

**Inclusion criteria**

- Diagnosis of treatment-resistant epilepsy of any syndrome, in which seizures are detectable in scalp EEG with two electrodes
- Between the ages of 18 – 80
- Experiencing >20 seizures per year according to seizure diary

**Exclusion criteria**

- Established current diagnosis of psychogenic non-epileptic attacks (dissociative seizures)
- Frequent vigorous involuntary movements (eg. chorea, athetosis) or frequent parasomnias with major motor components (eg. sleep walking, night terrors)
- Inability to comply with the trial procedure, such as cognitive or behavioral problems
- Inability to give informed consent
- History or evidence of: Severe cardiac disease (including Pacemaker and ICD-unit), Myocardial infarction, angina pectoris or other ischaemic heart disease, cardiac arrhythmia or any other heart failure
- History or evidence of recent (<1 year) stroke, transient ischaemic attack, carotid or vertebral artery stenosis or dissection, cerebral hemorrhage, or any other neurological disease deemed progressive or with high risk of recurrence, in the judgement of the CI
- Use of following drugs: Chemotherapeutic drugs of any kind, Methotrexate, Anticoagulation treatment, Immunosuppressant treatment, Third generation antipsychotic drugs (aripiprazole, quetiapine, clozapine, ziprasidone, paliperidone, risperidone, sertindole, amisulpride, olanzapine)
- Subjects under investigation or treatment of active cancer or cancer diagnosis within the past 5 years
- Subjects known with or suspected abuse of alcohol (defined as consumption of > 250g alcohol per week) or abuse of any other neuro-active substances
- Subjects involved in therapies with medical devices that deliver electrical energy into the area around the implant.
- Subjects at high risk of surgical complications, such as active systemic infection and hemorrhagic disease.
- Subjects who are allergic to the local anesthetics used during implantation.
- Subjects whose safety blood measurements (full blood count, U&E, clotting) are significantly out of range in the judgement of the CI.
- Female subjects of childbearing potential who are pregnant or intend to become pregnant or are not using adequate contraceptive methods throughout the study
- Subjects who have an infection at the site of device implantation.
- Subjects who operate MRI scanners or are planned to have an MRI scan within the next year.
- Subjects with profession/hobby that includes activity imposing extreme pressure variations (e.g. diving or parachute jumping). NB: diving/snorkeling is allowed to 5 meters of depth.
- Subjects with profession/hobby that includes activity imposing an unacceptable risk for trauma against the device or the site of implantation (e.g. martial arts or boxing).

### 2. Study Procedures Schematic

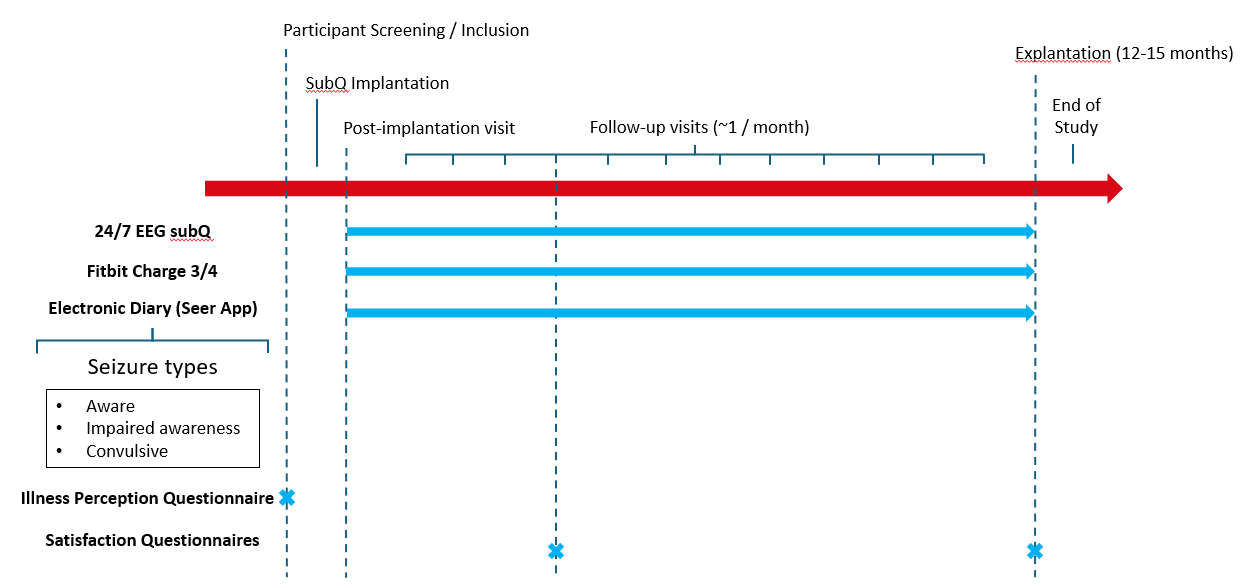

### 4. Components of the UNEEG 24/7 EEG^TM^ SubQ System

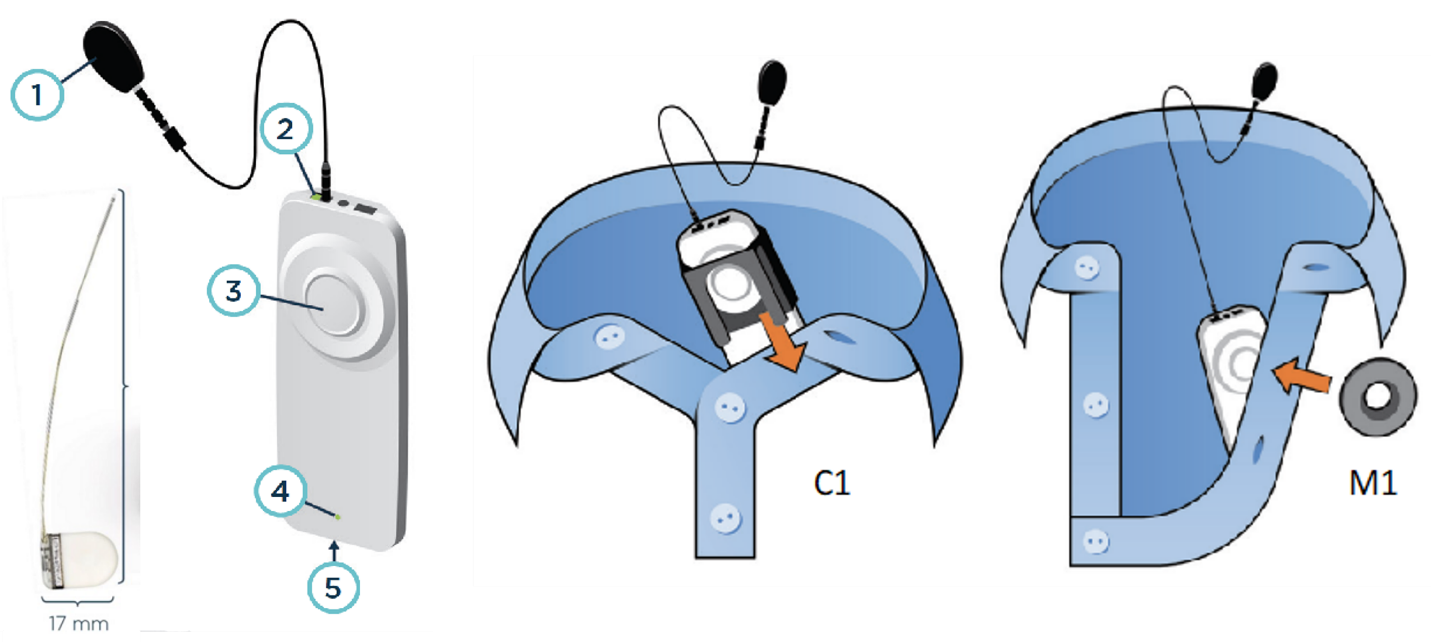

Left: components of the UNEEG 24/7 EEG™ SubQ system. The implant, consisting of an electrode and its attached housing, is implanted entirely subcutaneously. The device, that enables recording of the data and powers the implant, consists of several parts: 1) external cable, 2) connection light indicating if the device is connected to the implant, 3) power button, 4) charging light and 5) charger port. Right: the device attaches to the clothes via a plastic clip (C1) or a disc-shaped magnet (M1).

### 5. Data Quality Protocol Recording

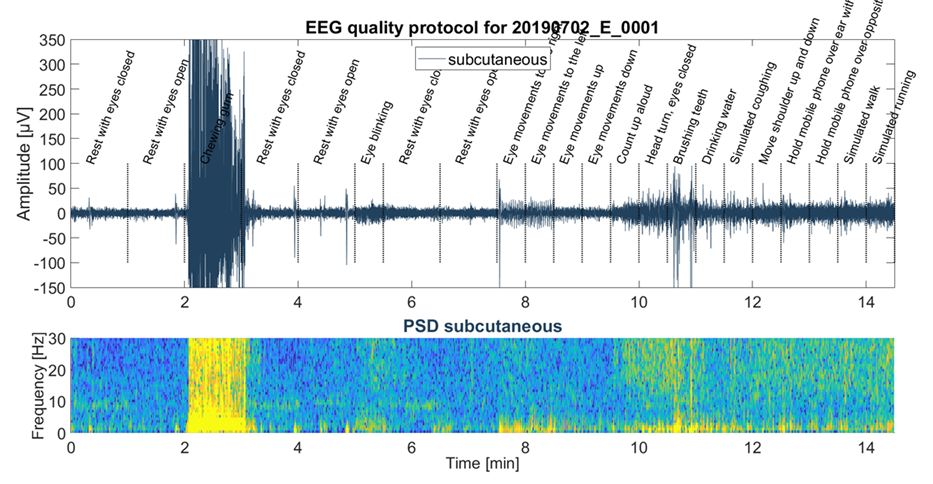

Example of a data quality protocol recording, done at the beginning of the study, showing the raw sqEEG of a single channel (top) and corresponding time-frequency plot (bottom panel), during different simulated daily life activities.

### 6. Baseline Brief Illness Perception Questionnaire

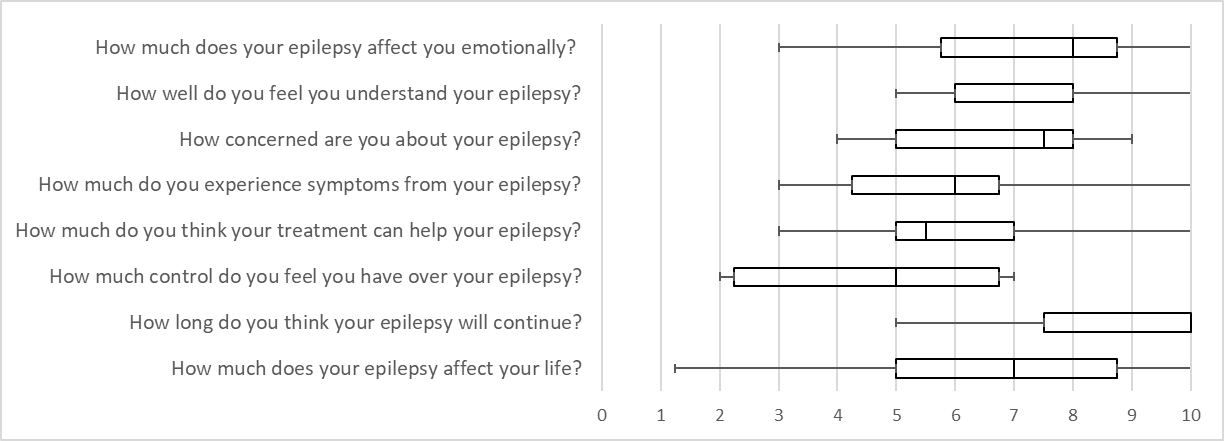

Brief Illness Perception Questionnaire at study baseline for all implanted patients (n=12). Boxplots of Likert Scale responses to each item.

### 7. Subject Satisfaction – Participant Comments

Examples on subject comments on acceptability questionnaires.

“Since having the implant, I forget it is there to be honest”, “Wouldn’t have thought it was there” and “No discomfort at any point”. “I am totally used to the device now. It took some getting used to and was a little uncomfortable at times - this was brief though.” “No discomfort experienced from the implant. I could occasionally feel it moving around under the skin but it wasn't harmful or unpleasant.”

Quotes from the same subject at 3 months post-implantion and 15 months post implantation, respectively:

“Once the usage of the device had been explained it was fairly straightforward.” (3 months post-implantation)

“After the initial explanation it was all fairly straightforward” (15 months post implantation)

“The SubQ was easy to use. I was shown how to set-up and use it within 20 mins and have been able to travel abroad with it since. I have had no issues that haven't been solved without the help of the troubleshoot guide” (3 months post-implant)

“The SUBQ interface is plug and play. It is as easy to use as most devices I have in my home” at end of study. ” (15 months post implantation)

“The research into the illness / condition is very important and I feel if I want to to help myself I have to take part. Once getting used to using the system it became easy so I feel other patients would soon get used to it. The system wasn't cumbersome but I had to be aware of it in case I dislodged it (i.e. learning to sleep on one side)” (3 months post-implantation)

“Trying to change clothes without having the device dangling was occasionally difficult. The clip would occasionally come undone and leave the device dangling. The stickers would occasionally loose effectiveness during the night and cause a loud noise to wake me up. The lack of water resistance allowed for occasions when I would have a seizure in the shower without the device to record it”. ” (15 months post implantation)

“No other comments. The booklet is helpful to read.” (3 months post-implantation)

“On the device itself, I would say – change the clip, you want it to be more flexible, more springy, less static, because if you wear a cardigan, then you expand it, then when you wear a tshirt it doesn’t stay close together anymore.” (15 months post-implantation)

### 8. UNEEG EpiSight Viewer Software

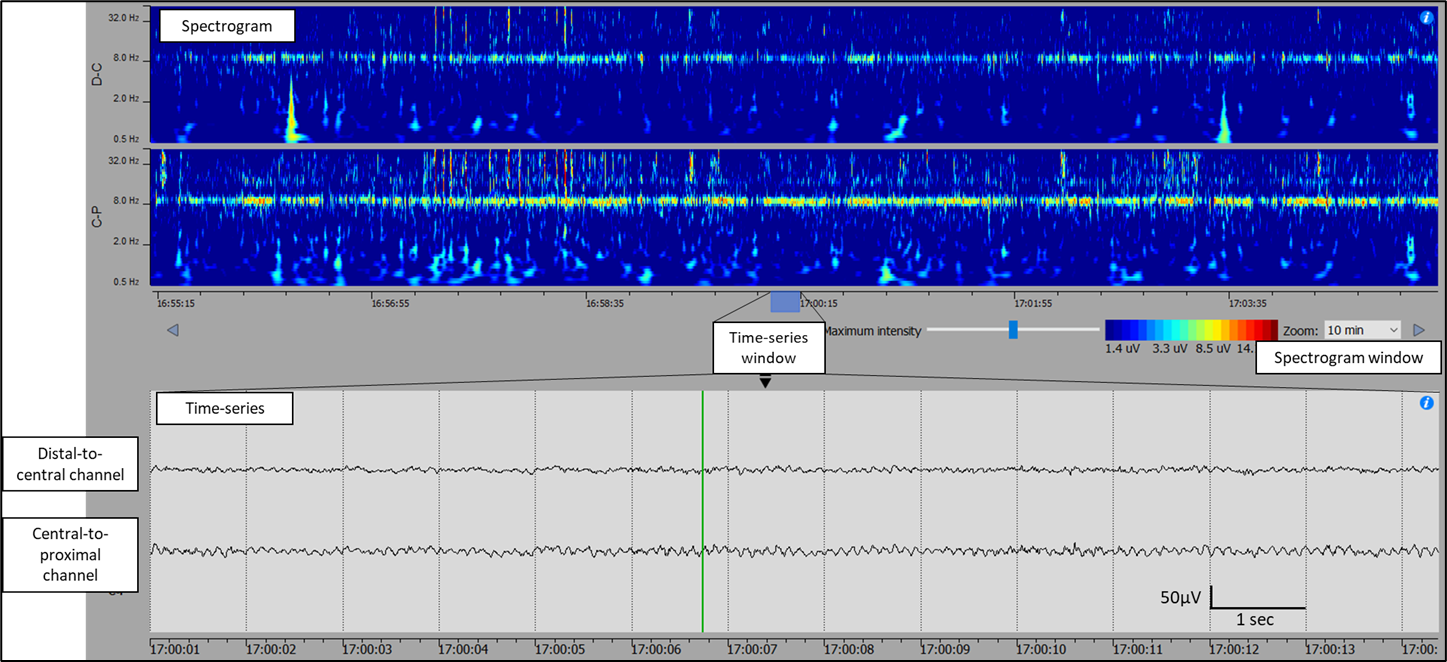

Supplementary Figure 5 – Example screenshot of UNEEG^TM^ Episight Viewer, showing 10-minute spectrogram plots at the top and raw time-series at the bottom.

### 9. Characterizing seizure types with sqEEG

A preliminary analysis was performed to assess the degree of inter-rater agreement of different seizure subtypes based on sqEEG recordings. We analysed 322 seizures from 15 patients with treatment-resistant epilepsy, comprising six patients from Weisdorf et al.^1^ and nine recordings from this study. Two independent raters (P.F.V. and C.S.) classified, based on visual interpretation, seizures into three subtypes:

1) Non-convulsive seizure with no / dubious clinical signs or artefacts
2) Non-convulsive seizure with clear clinical signs based on EMG/movement artefact (e.g. tonic, clonic or other stereotyped muscle artefact, or arousal)
3) Convulsive seizure

Results: Interrater agreement (Cohen’s k) for seizure subtype classification was 0.85 for all seizure types, and 1.0 for the differentiation between tonic-clonic and other seizures. Therefore, for this study, it was finally decided to split classification into non-convulsive (types 1 + 2) and convulsive (type 3) seizures.

|  | | Rater 2 | | |
| --- | --- | --- | --- | --- |
|  |  | Type 1 | Type 2 | Type 3 |
| Rater 1 | Type 1 | 80 | 0 | 0 |
|  | Type 2 | 22 | 205 | 0 |
|  | Type 3 | 0 | 0 | 15 |

### 10. Study Adverse Events

| ID | Type of AE | Severity | Relation to study participation? | Serious Adverse Event? | Outcome | Diagnosis |
| --- | --- | --- | --- | --- | --- | --- |
| SA | AE | Mild | Probable | No | Recovered without sequelae | Headache post-implantation |
| SA | ADE | Mild | Probable | No | Recovered without sequelae | Adverse device effect. Faulty device into error mode. Faulty implant. |
| SB | ADE | Mild | Probable | No | Recovered without sequelae | Adverse device effect. Faulty device into error mode. Faulty implant. |
| SC | AE | Mild | Unlikely | No | Recovered without sequelae | Headache |
| SF | AE | Mild | Probable | No | Recovered without sequelae | Headache |
| SF | AE | Moderate | Unlikely | No | Recovered without sequelae | Prolonged tonic-clonic seizure - assisted at home by ambulance crew |
| S10 | AE | Mild | Probable | No | Recovered without sequelae | Headache |
| S10 | AE | Severe | Probable | Yes - Hospitalisation | Recovered without sequelae | Electrode tip protrusion through the scalp requiring device explantation |
| SH | AE | Mild | Unlikely | No | Recovered without sequelae | Fall |
| S13 | AE | Severe | Unlikely | Yes - Hospitalisation | Recovered without sequelae | Community Acquired Pneumonia |
| S13 | AE | Moderate | Possible | No | Recovered without sequelae | Headache |
| SJ | AE | Mild | Unlikely | No | Recovered without sequelae | Electrode tip protrusion through the scalp during explantation and reimplantation procedure |

### 11. Device Deficiencies

| Subject | Device type | Nature of the deficiency | Outcome / Solution | Affected data collection? |
| --- | --- | --- | --- | --- |
| SA | External Logger | Error mode (likely related to implant malfunction) | Replacement, rebooting (and later reimplantation) | Yes |
| SA | External Logger | Error mode (likely related to implant malfunction) | Replacement, rebooting (and later reimplantation) | Yes |
| SA | External Logger | Error mode (likely related to implant malfunction) | Replacement, rebooting (and later reimplantation) | Yes |
| SA | External Logger | Error mode (likely related to implant malfunction) | Replacement, rebooting (and later reimplantation) | Yes |
| SA | External Logger | Error mode (likely related to implant malfunction) | Replacement, rebooting (and later reimplantation) | Yes |
| SA | External Logger | Error mode (likely related to implant malfunction) | Replacement, rebooting (and later reimplantation) | Yes |
| SA | External Logger | Error mode (likely related to implant malfunction) | Replacement, rebooting (and later reimplantation) | Yes |
| SA | External Logger | Error mode (likely related to implant malfunction) | Replacement, rebooting (and later reimplantation) | Yes |
| SA | External Logger | Error mode (likely related to implant malfunction) | Replacement, rebooting (and later reimplantation) | Yes |
| SA2 | External Logger | Disconnecting more frequently than usual, patient reluctant to wear at night to avoid being woken up by disconnection alarm | Replacement | Yes |
| SA2 | External Logger | Disconnecting more frequently than usual, patient reluctant to wear at night to avoid being woken up by disconnection alarm | Replacement | Yes |
| SA2 | External Logger | Disconnecting frequently | Replacement | Yes |
| SA2 | External Logger | Error mode (unclear cause) | Rebooting and replacement | Yes |
| SA2 | External Logger | Damaged cable (chewed by mouse) | Replacement | Yes |
| SB | External Logger | Error mode (likely related to implant malfunction) | Explantation, study drop-out | Yes |
| SB | External Logger | Error mode (likely related to implant malfunction) | Explantation, study drop-out | Yes |
| SD | External Logger | Wron out cable, kept disconnecting | Replacement | Yes |
| SD | External Logger | Wrong timestamp (timestamped to 1/1/1900) | Rebooting and replacement | No |
| SD | External Logger | Error mode (unclear cause) | Rebooting and replacement | Yes |
| SE | External Logger | Disconnecting frequently | Replacement | Yes |
| SF | External Logger | Disconnecting frequently | Replacement | Yes |
| SF | External Logger | Volume drastically reduced (noticed after a trip to the dentist) | Replacement | Yes |
| SH | External Logger | The hook lost its grip to clothes | Replacement | Yes |
| SJ | External Logger | Disconnecting frequently | Replacement | Yes |
| SA2 | Fitbit | Fitbit stopped working | Replacement | Yes |
| SA2 | Fitbit | Broken strap | Replacement | Yes |
| SF | Fitbit | Broken strap | Replacement | Yes |
| SD | Laptop | Wrong timezone | Replacement of laptop, reparsement of data | No |
| SE | Laptop | Wrong timezone | Replacement of laptop, reparsement of data | No |
| SF | USB | Data not received via post | Backup data retrieved from laptop | No |
| SA | Implant | Implant Malfunction | Reimplantation | Yes |
| SB | Implant | Implant Malfunction | Explantation, study drop-out | Yes |

### 12. Circadian Adherence Plots

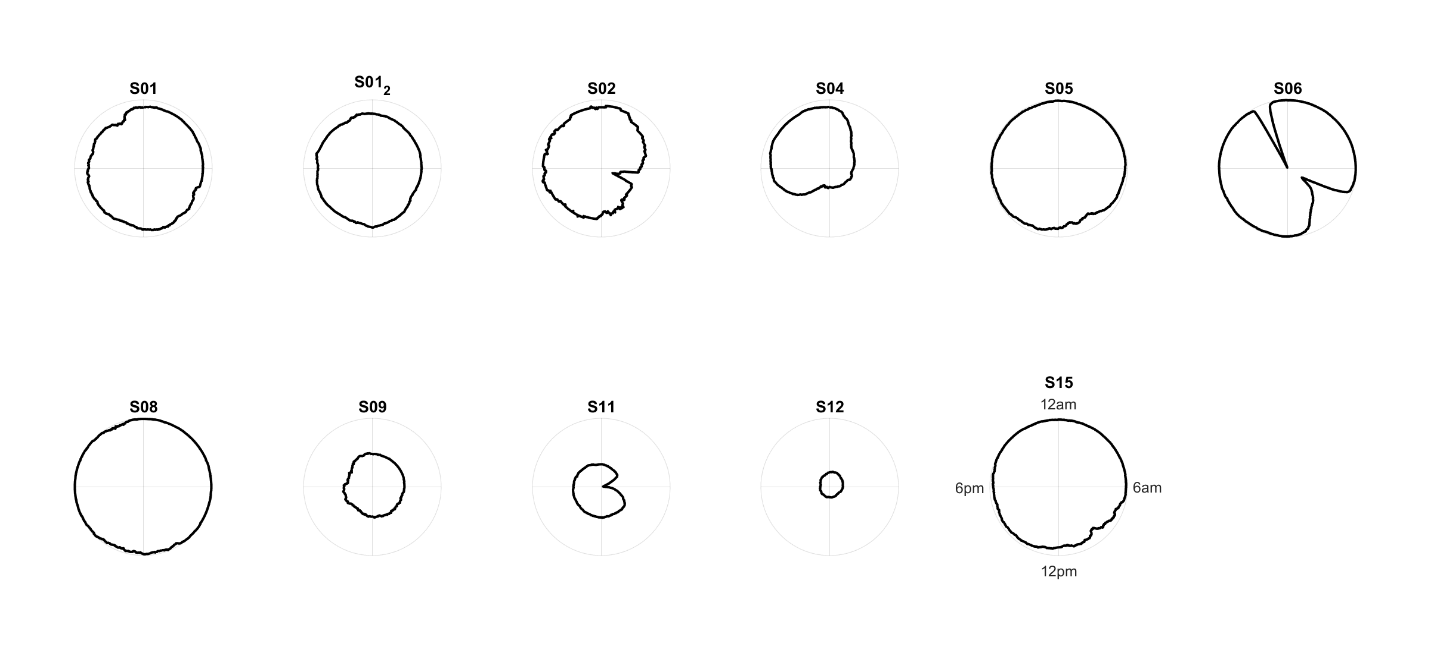

### 13. Weekday effect on adherence to sqEEG

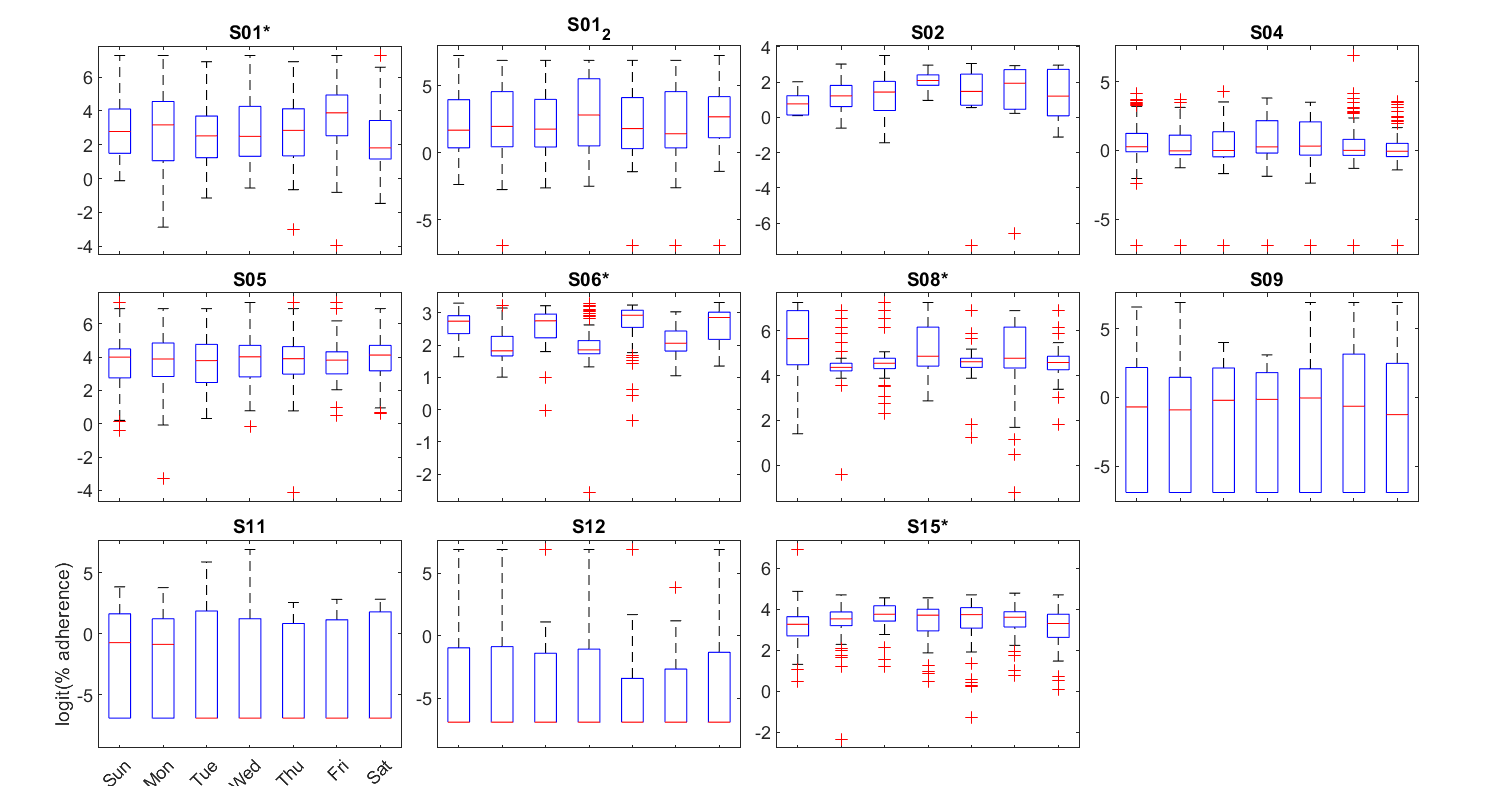

Boxplots of percentage adherence per participant, per day of the week.

*Significant weekday effect (p<0.05 on Kruskall-Wallis tests).

### 14. Longterm trend in adherence – individual Linear Models

| Subject | intercept | slope | p-value | Adjusted R-Squared |
| --- | --- | --- | --- | --- |
| SA | 0.011 | 0.011 | <0.001 | 0.124 |
| SA2 | -0.003 | -0.003 | <0.001 | 0.032 |
| SB | -0.054 | -0.054 | 0.017 | 0.102 |
| SC | 0.001 | 0.001 | 0.192 | 0.001 |
| SD | -0.003 | -0.003 | <0.001 | 0.049 |
| SE | 0.000 | 0.000 | 0.841 | -0.002 |
| SF | 0.001 | 0.001 | 0.002 | 0.020 |
| SG | -0.045 | -0.045 | <0.001 | 0.585 |
| SH | -0.018 | -0.018 | <0.001 | 0.291 |
| SI | 0.003 | 0.003 | 0.239 | 0.001 |
| SJ | -0.001 | -0.001 | 0.003 | 0.019 |

### 15. Example seizure patterns

SA – nonconvulsive

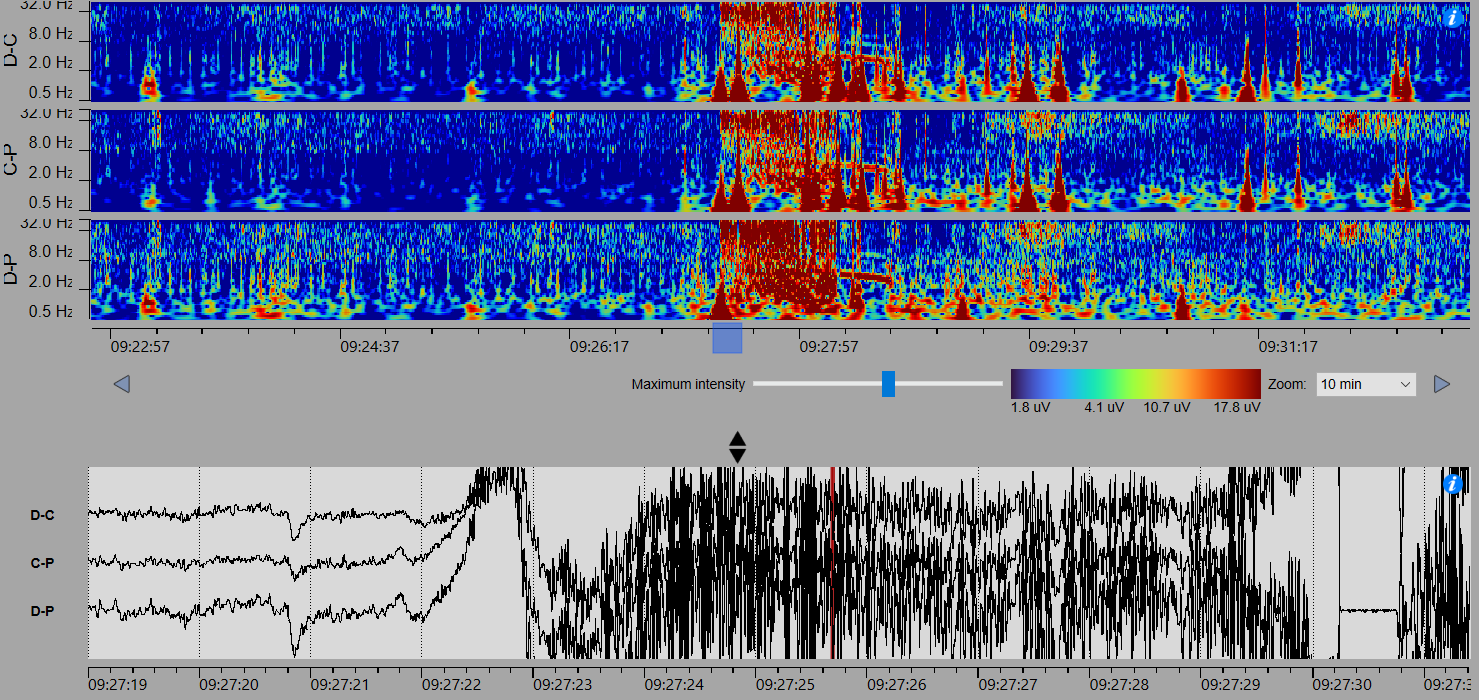

SA2 – nonconvulsive

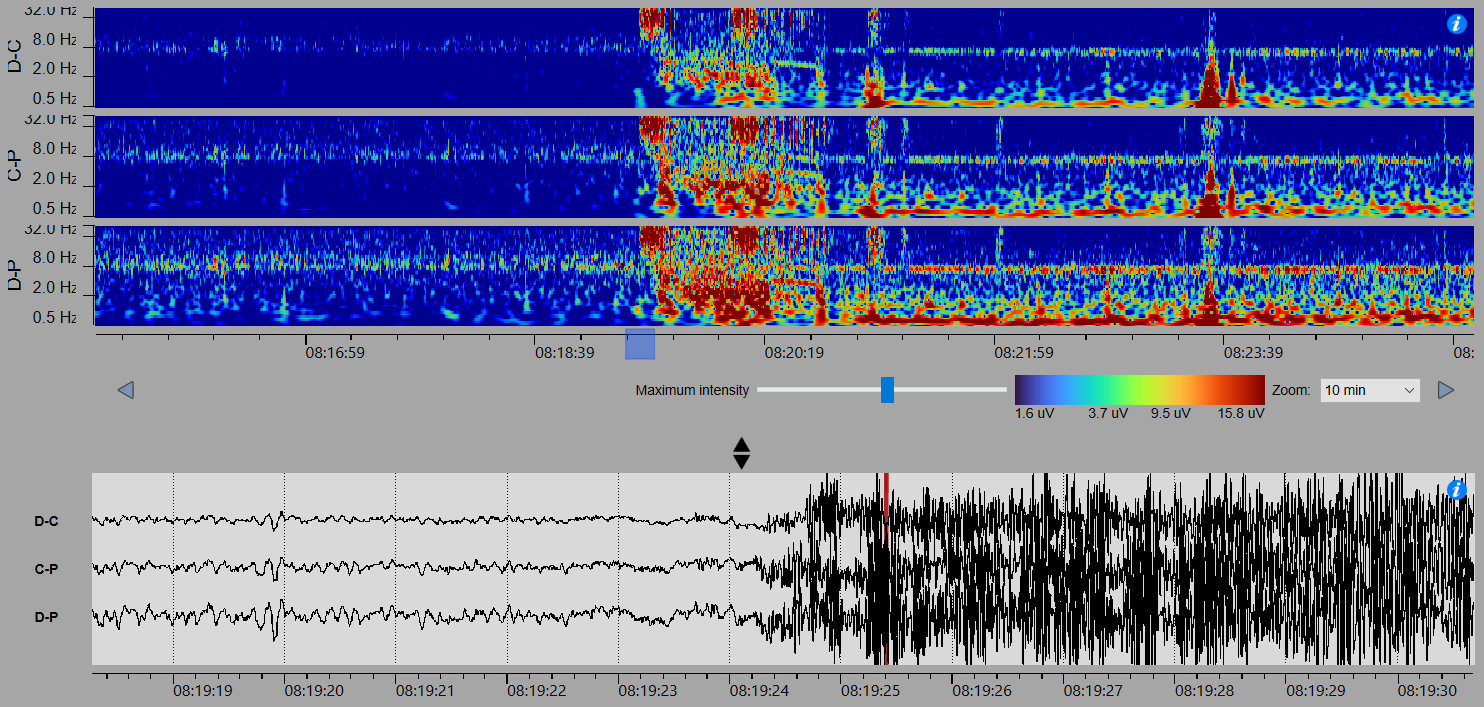

SB - nonconvulsive

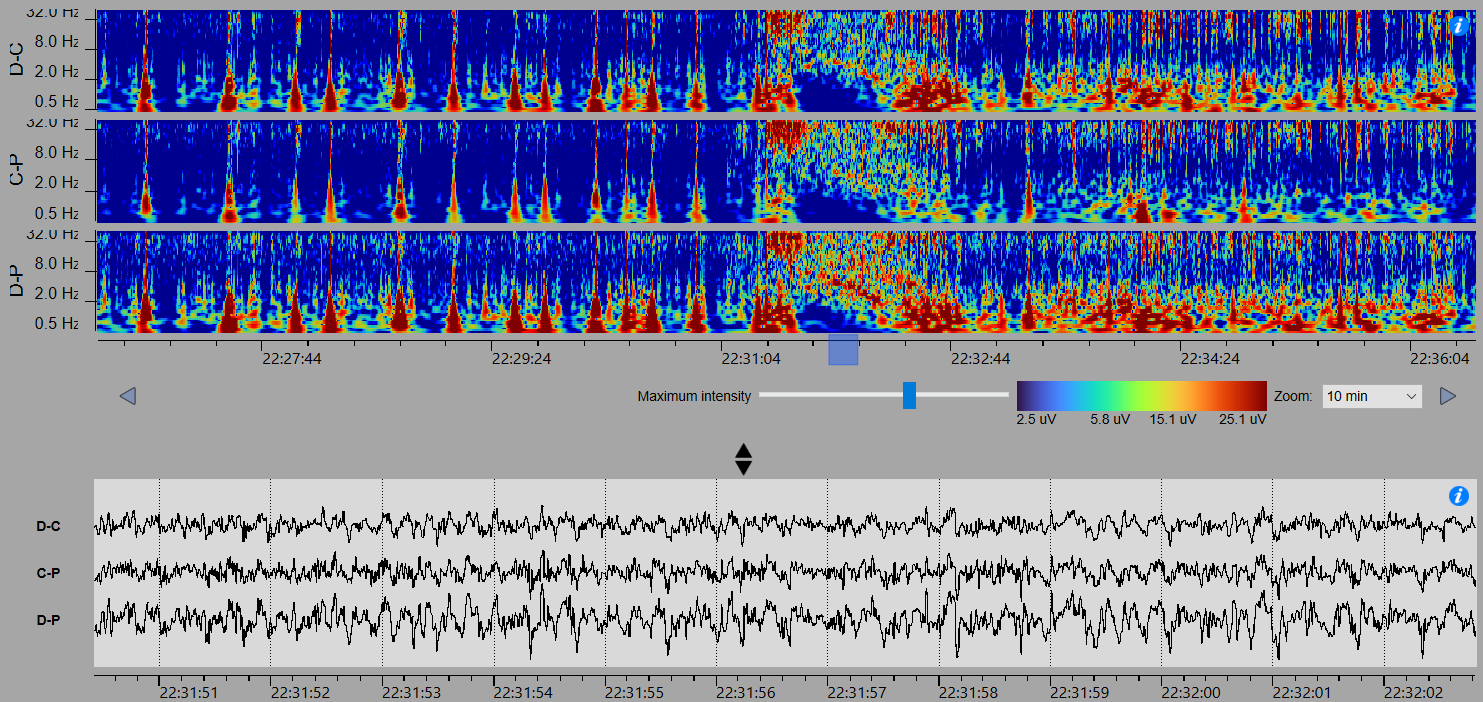

SC - nonconvulsive

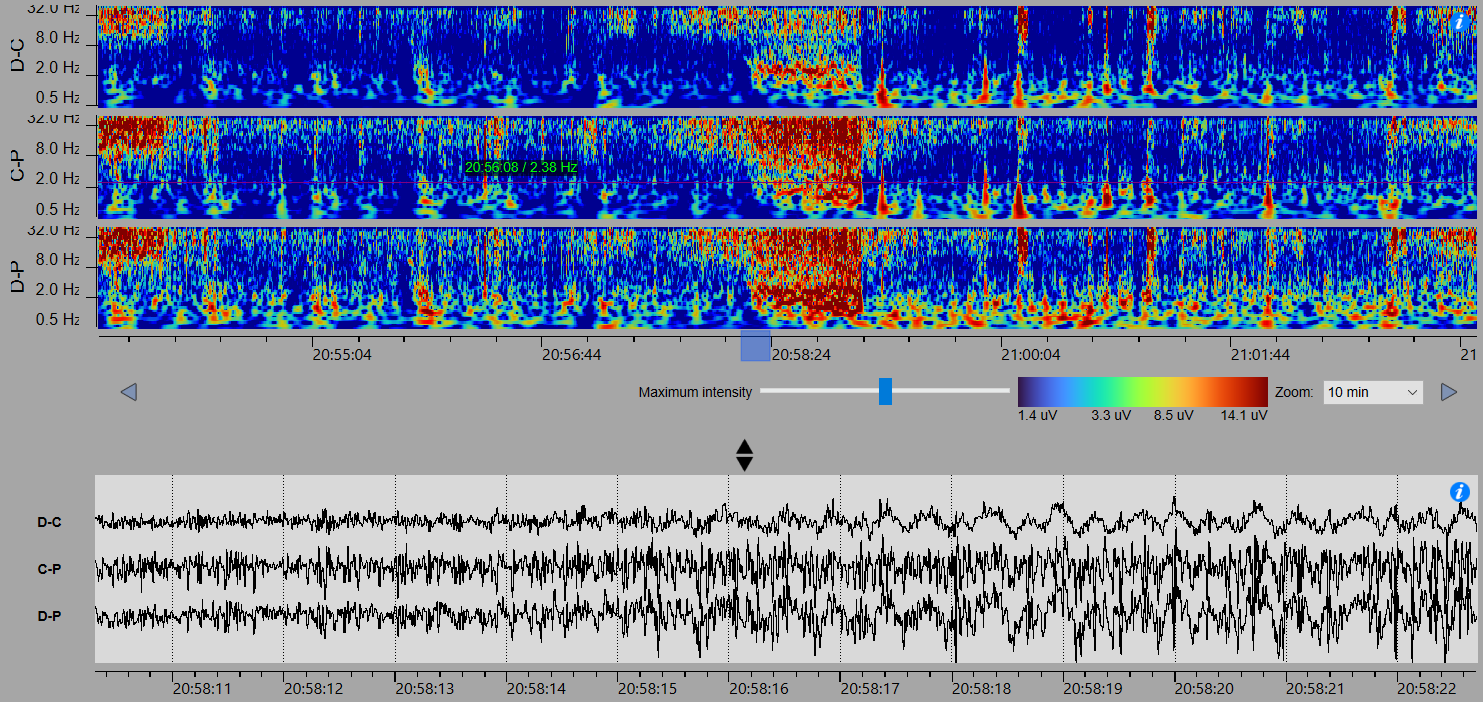

SD - nonconvulsive

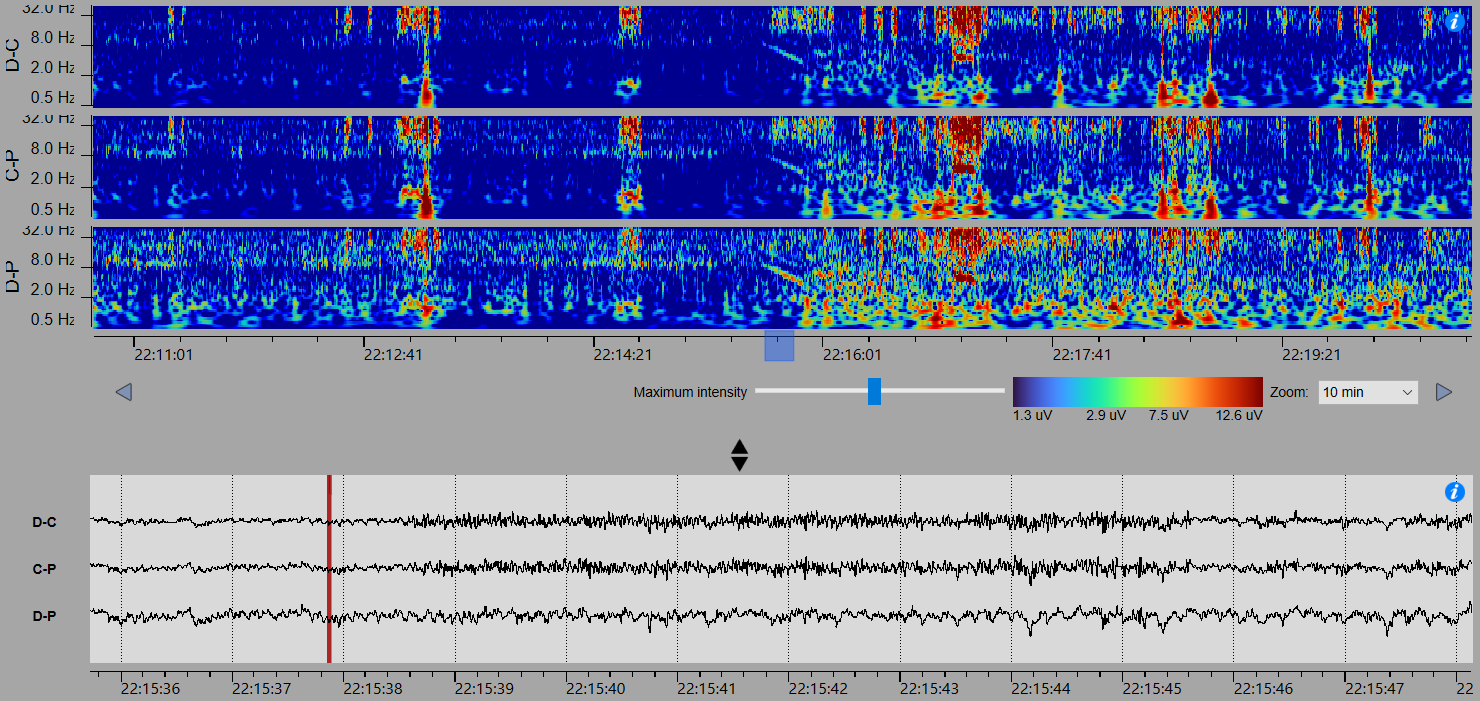

SE - nonconvulsive

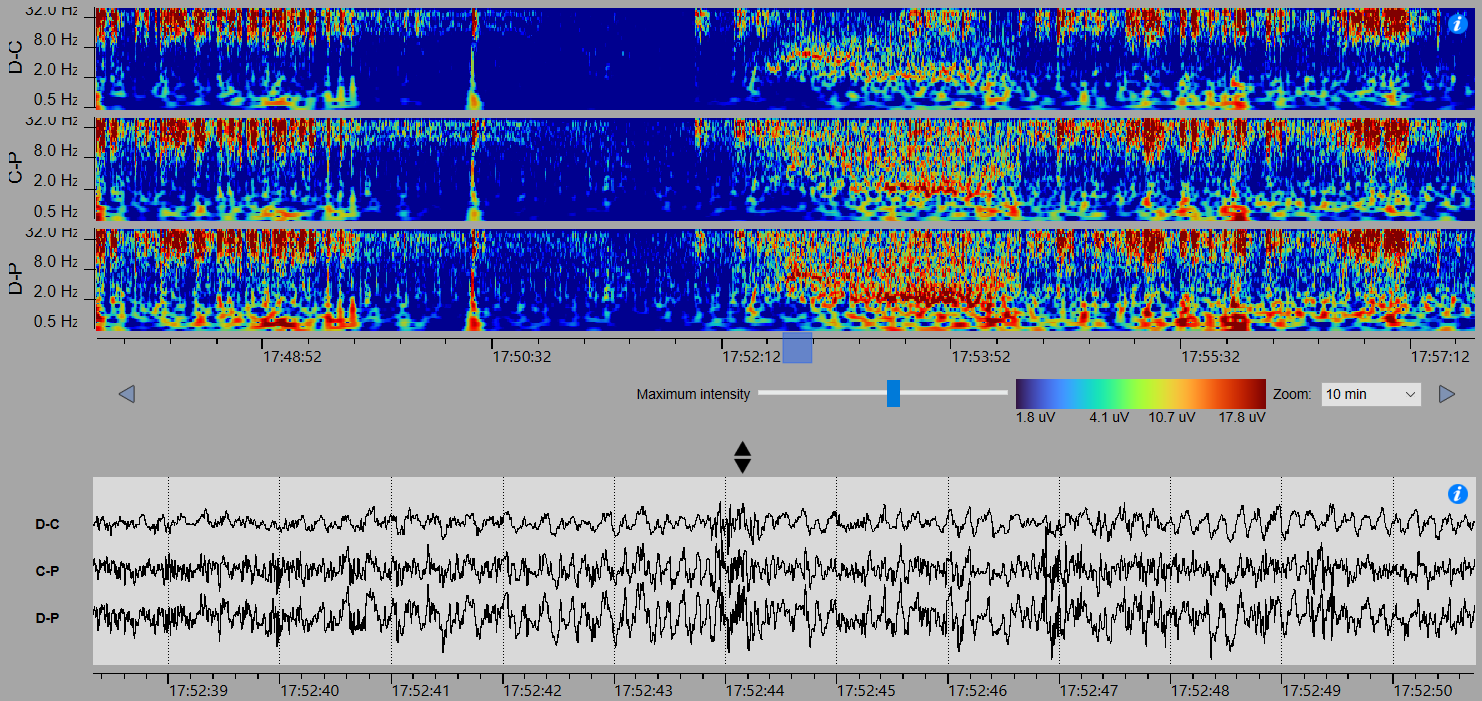

SF - nonconvulsive

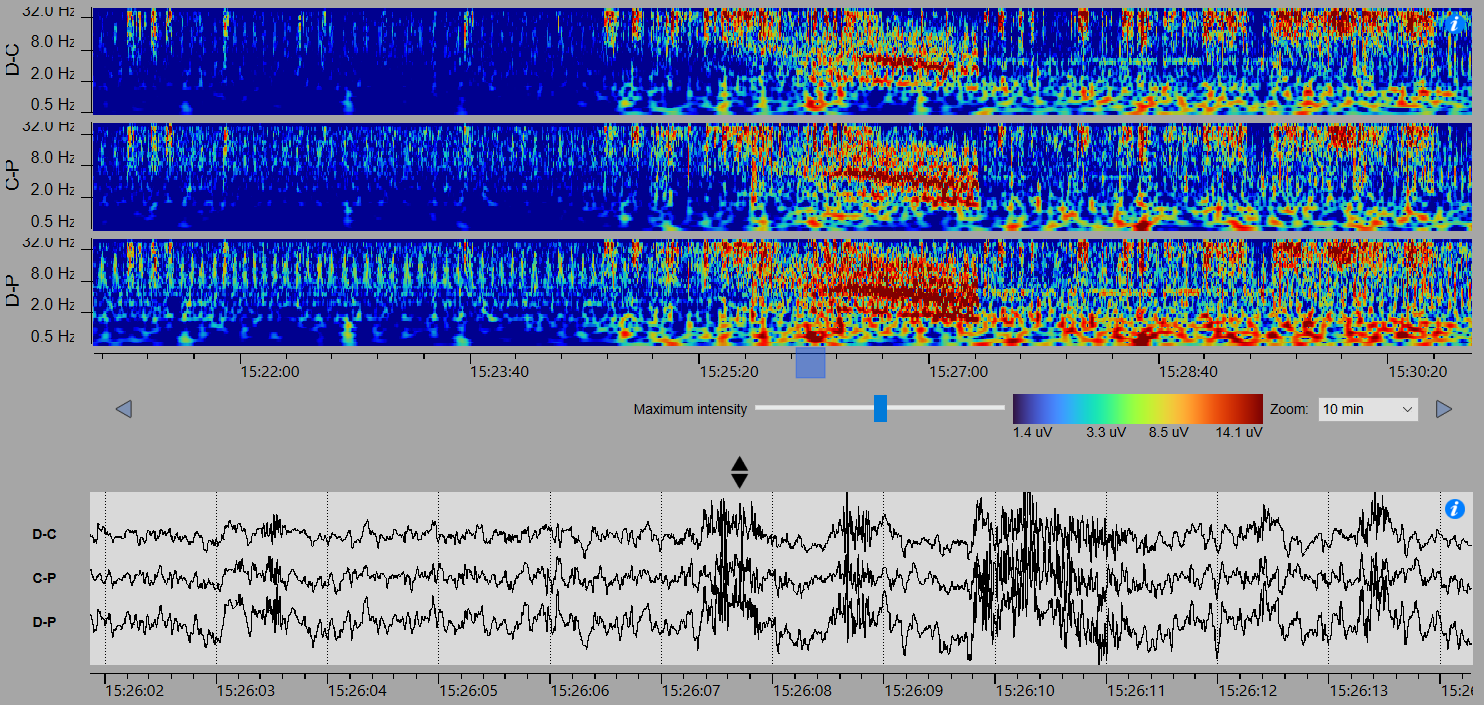

SF - Convulsive

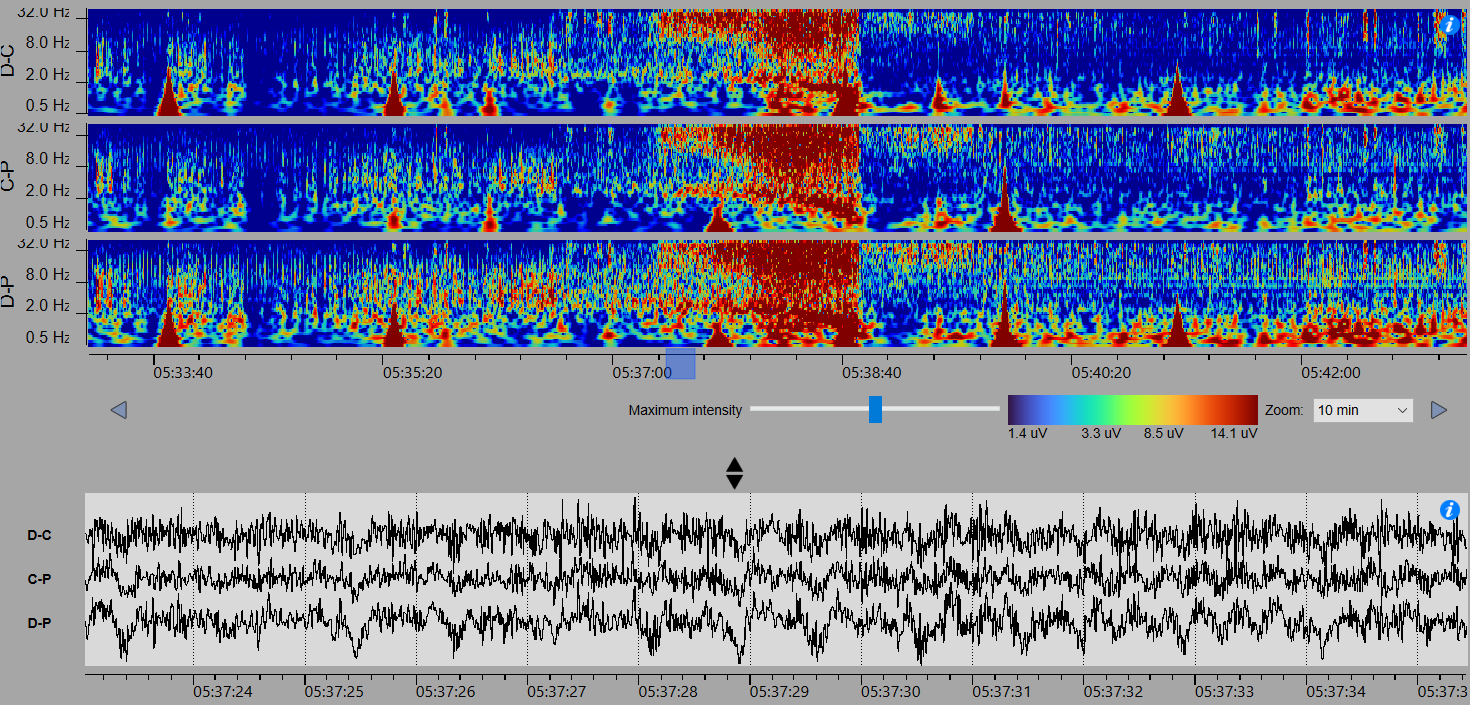

SG - convulsive

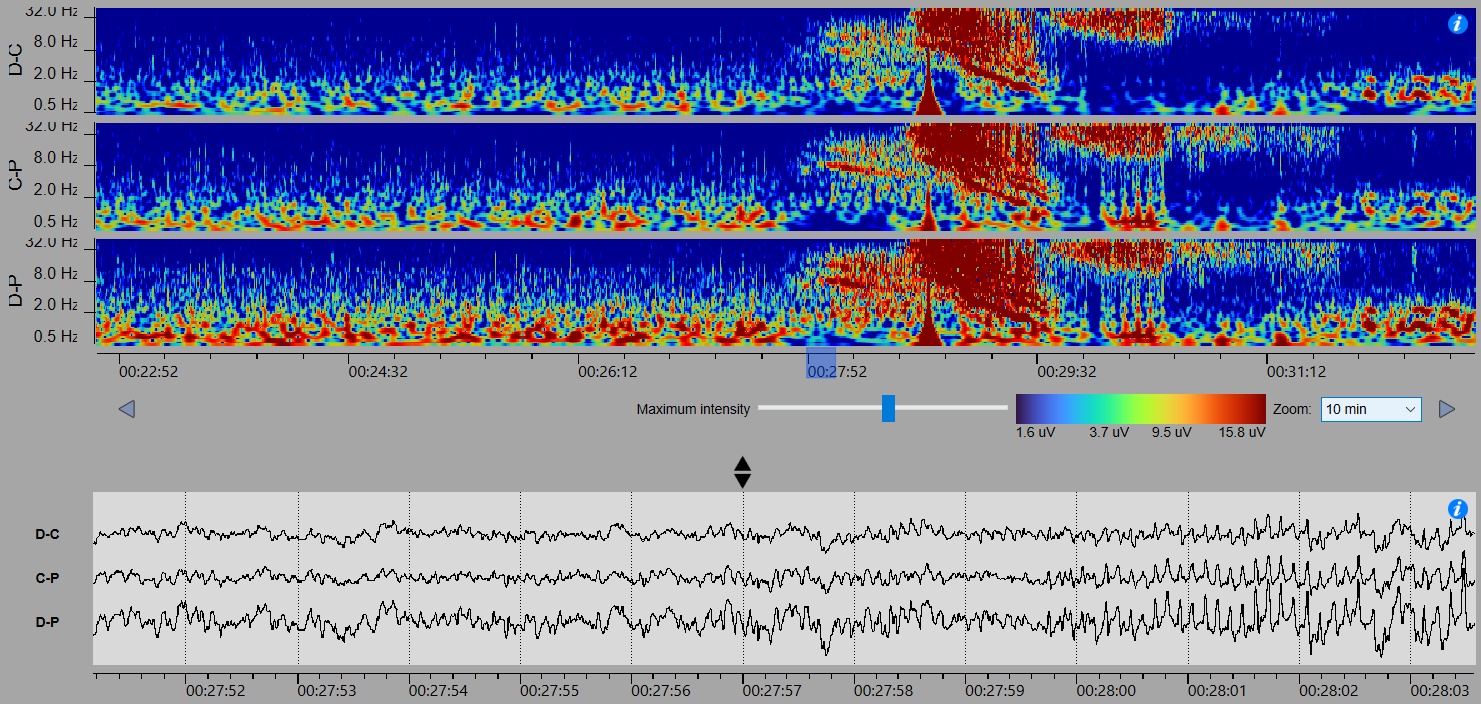

SG - nonconvulsive

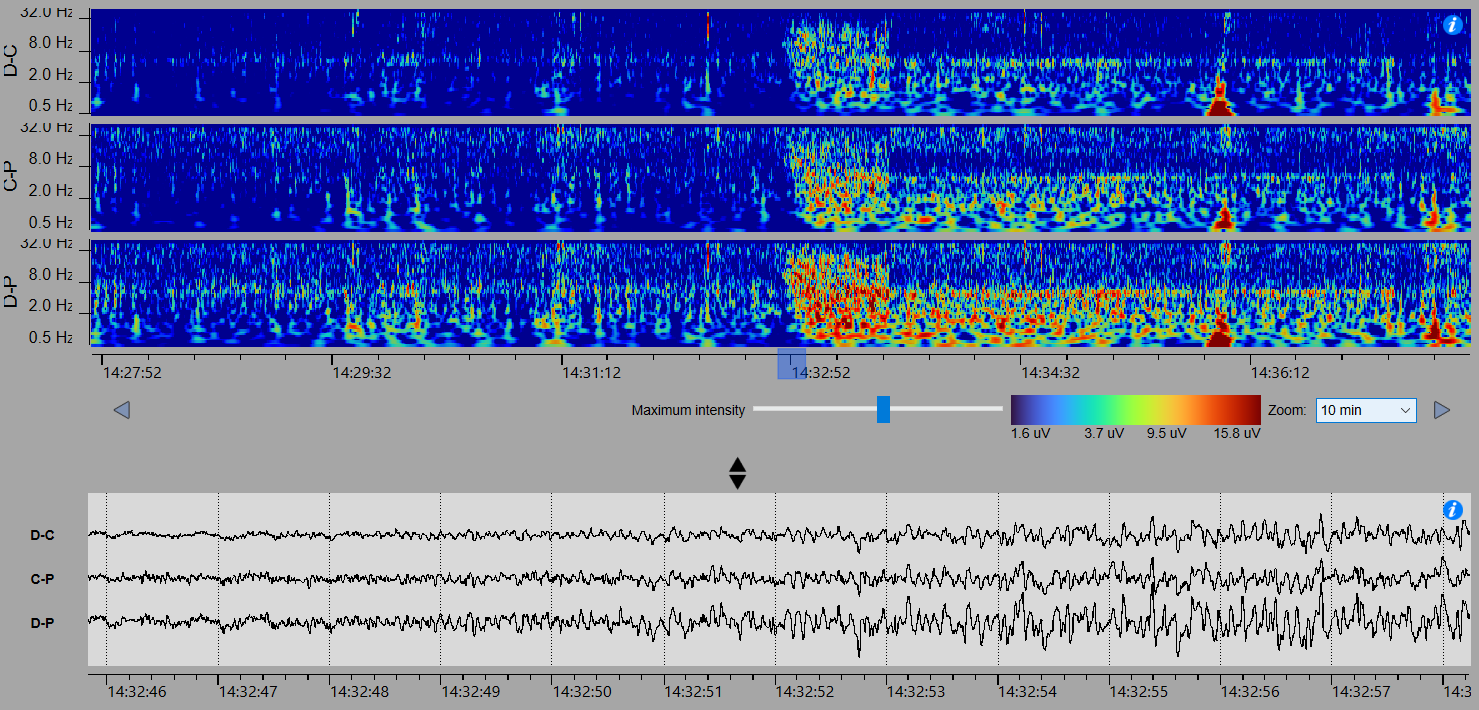

SH – nonconvulsive

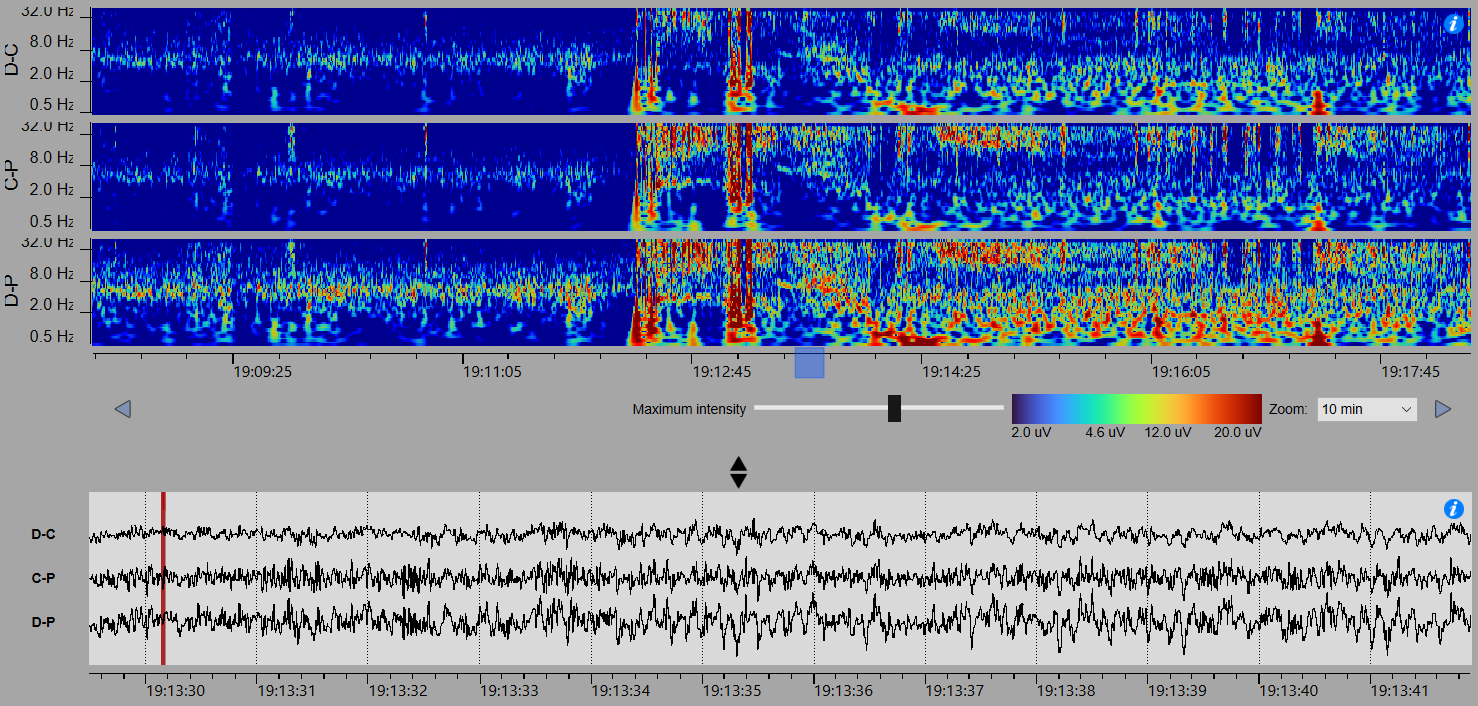

SJ - nonconvulsive

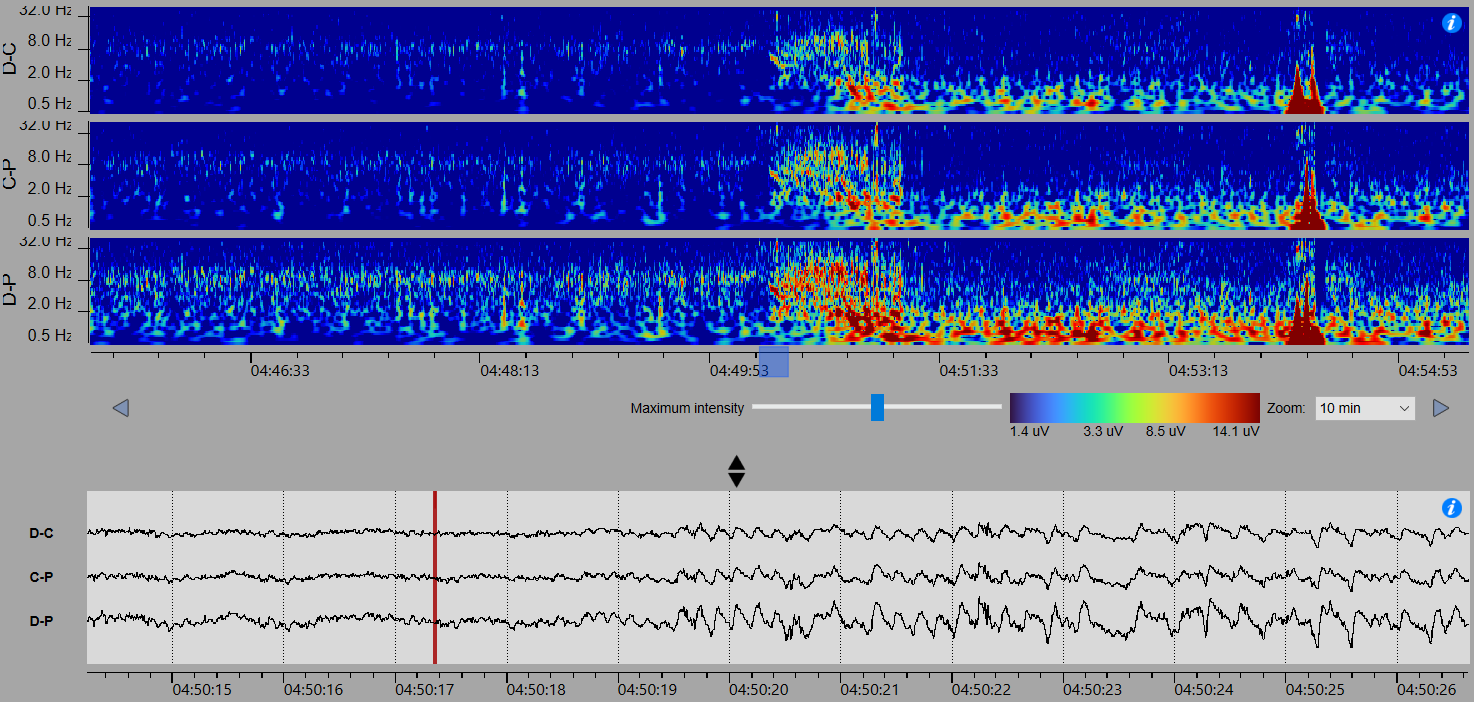

### 16. Seizure Detection Algorithm Performance – Additional Results

|  |  |  |  |  | Detected seizures (n=470) | | Undetected seizures (n=284) | | | | |  |
| --- | --- | --- | --- | --- | --- | --- | --- | --- | --- | --- | --- | --- |
| ID | N. seizures (n) | N. automatic detections (n) | N. diary events (n) | Diary events whilst recording sqEEG | Total (n) | Detected on random 10% review periods (n) | Identified on random 10% review (n) | Identified on review of diary events (n) | Identified on full review only (n) | Identified by chance (n) | Total (n) | Estimated % of recording reviewed^#^ |
| SA* | 33 | 4768 | 24 | 22 | 33 | 7 | 0 | 0 | N/A | 0 | 0 | 18% |
| SA2 | 92 | 1244 | 41 | 37 | 76 | 9 | 1 | 12 | N/A | 3 | 16 | 12% |
| SB | 36 | 127 | 28 | 21 | 21 | 3 | 4 | 15 | 0 | 0 | 15 | 100% |
| SC | 56 | 3468 | 61 | 37 | 52 | 7 | 0 | 3 | N/A | 1 | 4 | 13% |
| SD | 54 | 1923 | 49 | 47 | 7 | 0 | 2 | 33 | 13 | 0 | 47 | 100% |
| SE | 54 | 984 | 55 | 52 | 54 | 3 | 0 | 0 | N/A | 0 | 0 | 11% |
| SF | 132 | 7047 | 136 | 129 | 41 | 3 | 9 | 64 | 24 | 0 | 91 | 100% |
| SG | 203 | 463 | 133 | 115 | 114 | 20 | 2 | 62 | 25 | 0 | 89 | 100% |
| SH | 31 | 2404 | 2 | 2 | 17 | 3 | 9 | 1 | 13 | 0 | 14 | 100% |
| SI | 0 | 193 | 24 | 6 | 0 | 0 | 0 | 0 | 0 | 0 | 0 | 100% |
| SJ | 63 | 1269 | 39 | 38 | 55 | 7 | 1 | 7 | N/A | 0 | 8 | 12% |

*initial version (v1.11) of the seizure detector

^#^10% of recordings + 5 minutes around automated event markers + 2 hours around diary periods, or full review
